# Combined physical and cognitive training for older adults with and without cognitive impairment: A systematic review and network meta-analysis of randomized controlled trials

**DOI:** 10.1101/2020.08.08.20170654

**Authors:** Hanna Malmberg Gavelin, Christopher Dong, Ruth Minkov, Alex Bahar-Fuchs, Kathryn A Ellis, Nicola T Lautenschlager, Maddison L Mellow, Alexandra T Wade, Ashleigh E Smith, Carsten Finke, Stephan Krohn, Amit Lampit

**Author notes:** Correspondence concerning this article should be addressed to Amit Lampit,.

## Abstract

Combining physical exercise with cognitive training is a popular intervention in dementia prevention trials and guidelines. However, it remains unclear what combination strategies are most beneficial for cognitive and physical outcomes. We aimed to compare the efficacy of the three main types of combination strategies (simultaneous, sequential or exergaming) to either intervention alone or control in older adults. Randomized controlled trials of combined cognitive and physical training were included in multivariate and network meta-analyses. In cognitively healthy older adults and mild cognitive impairment, the effect of any combined intervention relative to control was small and statistically significant for overall cognitive (*k*=41, Hedges’ *g* = 0.22, 95% CI 0.14 to 0.30) and physical function (*k*=32, *g* = 0.25, 95% CI 0.13 to 0.37). Simultaneous training was the most efficacious approach for cognition, followed by sequential combinations and cognitive training alone, and significantly better than physical exercise. For physical outcomes, simultaneous and sequential training showed comparable efficacy as exercise alone and significantly exceeded all other control conditions. Exergaming ranked low for both outcomes. Our findings suggest that simultaneously and sequentially combined interventions are efficacious for promoting cognitive alongside physical health in older adults, and therefore should be preferred over implementation of single-domain training.

## 1. Introduction

As population ageing increases globally, cognitive impairment and dementia have become a leading cause of disability (World Health Organization and Alzehimer’s Disease International, 2012). Modifiable risk factors such as physical and cognitive inactivity, depression, social isolation and poor cardiovascular health have been estimated to account for about 40% of the risk for developing dementia across the lifespan (Livingston et al., 2020b, a) and are recognized as important targets for non-pharmacological interventions aiming to prevent or delay cognitive decline. Multimodal intervention strategies have received particular interest, with over a dozen ongoing trials testing the assumption that targeting multiple risk factors would lead to additive or even synergetic effects on preserving functional independence in older age (Kivipelto et al., 2020).

Recent systematic reviews have supported the efficacy of cognitive training (Gavelin et al., 2020; Hill et al., 2017; Lampit et al., 2014; Leung et al., 2015) and physical exercise (Falck et al., 2019; Northey et al., 2018) on cognitive performance in older adults with and without cognitive impairment. These interventions have also been recommended in clinical practice guidelines (e.g., for mild cognitive impairment; Petersen et al., 2018) and, increasingly, the possibility of combining physical exercise with cognitively challenging activities has been highlighted as a way to maintain both cognitive and physical health (Yang et al., 2019).

Previous systematic reviews and meta-analyses have reported cognitive benefits following combined cognitive and physical interventions in older adults with or without cognitive impairment (Gheysen et al., 2018; Karssemeijer et al., 2017; Stanmore et al., 2017; Zhu et al., 2016), suggesting efficacy over and above physical exercise alone, and comparable to that of cognitive training. However, these meta-analyses mixed older with younger populations (Stanmore et al., 2017) and randomized controlled trials (RCT) with other designs (Gheysen et al., 2018; Zhu et al., 2016) or included a combination of cognitive training and other cognition-oriented treatments, such as cognitive stimulation or non-specific cognitive activities (Karssemeijer et al., 2017). Furthermore, most meta-analyses have so far focused on cognitive outcomes (Gheysen et al., 2018; Stanmore et al., 2017; Zhu et al., 2016) and the efficacy of combined interventions on physical and everyday function in general, and compared to more traditional physical exercise interventions in particular, is not yet established.

A key outstanding question in the field is how the efficacy of combined physical and cognitive interventions is related to the mode of delivery. Combined interventions can be broadly divided into sequential or simultaneous designs. Sequential designs deliver the intervention modalities in separate sessions, usually during the same period (Ngandu et al., 2015) or, less commonly, on separate periods throughout the course of the intervention (Heffernan et al., 2019). Simultaneous interventions are usually based on either delivery of cognitive training while asking participants to exercise at the same time, or by using exergaming, i.e., physically active videogames that include cognitively challenging tasks (see definitions under *2.3.3. Types of interventions* below).

While previous meta-analyses have provided some indications that simultaneous training leads to larger effects on cognition than those of sequential designs (Gheysen et al., 2018; Zhu et al., 2016), results have so far been inconclusive. Importantly, this question is difficult to ascertain in pairwise meta-analysis, since the relative efficacy of combined interventions is likely to be influenced by the type of control group activity (i.e., cognitive training or physical exercise alone, active or passive control), which varies considerably across studies (Gheysen et al., 2018; Zhu et al., 2016). In this context, network meta-analysis can be used to compare efficacy estimates of the different delivery formats to single modality or other types of control conditions. Such an approach has been conducted in one previous systematic review (Bruderer-Hofstetter et al., 2018); however, due to the large number of treatment nodes and relatively small number of included studies, the precision of the effect estimates was limited and the quality of the evidence was rated as very low. Finally, the efficacy of exergaming in older adults is still unclear due to the limitations noted above as well as a recent re-analysis of Stanmore et al. (2017), suggesting considerably smaller effects and potential small study bias (Sala et al., 2019).

The aim of the current review is therefore twofold: (1) conduct pairwise meta-analysis to investigate the efficacy of combined cognitive and physical training interventions on cognitive, physical, psychosocial and functional outcomes in older adults across RCTs; and (2) apply network meta-analysis to compare and rank the efficacy of the three main types of combined intervention delivery formats (simultaneous, sequential and exergaming) on cognitive and physical function relative to either intervention alone or inert control conditions.

## 2. Method

### 2.1 Protocol and registration

This work adheres to the Preferred Reporting Items for Systematic reviews and Meta-Analyses (PRISMA) guidelines (Liberati et al., 2009) and PRISMA extension for network meta-analysis (Hutton et al., 2015) and was prospectively registered with PROSPERO (CRD42020143509).

### 2.2 Search strategy and study selection

We searched MEDLINE, Embase and PsycINFO from inception to 23 July 2019 to identify RCTs examining the effects of combined cognitive and physical training on cognitive, physical, psychosocial or functional outcomes (see Appendix A for the full search strategy). No restrictions on language or publication type were applied. Two independent reviewers (CD and MM or AW) conducted initial screening of titles and abstracts and assessed full-text versions of potentially relevant articles. Disagreements were resolved by a third reviewer (AL or HMG). The electronic search was complemented by hand-searching the references of included papers and previous reviews.

### 2.3 Eligibility criteria

#### 2.3.1 Types of studies

Published, peer-reviewed reports of RCTs investigating the effects of a combined cognitive and physical exercise intervention on one or more cognitive, physical, psychosocial or functional outcome in older adults were included. No restrictions on the type or size of randomized trials were applied in order to ensure that all relevant literature was included. The primary outcome was cognitive function; therefore, eligible studies needed to provide at least one untrained cognitive outcome. Studies were included if they compared a combined intervention with cognitive or physical training alone, a control intervention or a passive control group. Randomized crossover trials were included, but only the first treatment phase was considered for analysis to avoid the influence of potential carryover effects.

#### 2.3.2 Types of participants

Studies were included if they focused on older adults and had a mean age of 60 years or older. This included cognitively healthy older adults, people with subjective cognitive complaints, mild cognitive impairment (MCI), dementia or Parkinson’s disease, all are populations for which the efficacy of cognitive training or physical exercise have been evaluated in previous systematic reviews. Eligibility was confirmed by examining the study inclusion criteria and baseline characteristics of the sample.

#### 2.3.3 Types of interventions

Studies were included if they focused on interventions combining process-based cognitive training with structured physical exercise. Process-based cognitive training was defined as repeated practice on tasks targeting one or several cognitive domains, as opposed to explicit learning of strategies (Lustig et al., 2009). Physical exercise included any form of structured physical activity, such as aerobic exercise, strength or functional (i.e., gait or balance) training. Combined interventions could be delivered as (1) *simultaneous training*: cognitive training and physical exercise delivered concurrently in a dual-task format; (2) *sequential training*: cognitive training and physical exercise delivered in separate sessions, either on the same day or on different days; or (3) *exergaming*: physically active video games including cognitively challenging tasks. Exergaming interventions were included if the games placed cognitive demands, such as requiring attention, processing speed, planning or decision making, at a level commensurate with process-based cognitive training; pure exercise or sport games, such as yoga or balance exercises were excluded. To allow the inclusion of multicomponent strategies, interventions including other components such as psychoeducation, strategy-based cognitive training or diet were eligible as long as cognitive training and physical exercise were each provided for at least 20% of the total intervention time. Studies with interventions that comprised more than 20% of one intervention but less than 20% of the other (e.g., Ngandu et al., 2015) were excluded.

#### 2.3.4 Types of controls

Studies were included if they compared a combined intervention with physical exercise, cognitive training, a sham intervention (e.g. health education, relaxation, stretching or non-specific cognitive activities such as data entry on a computer) or a passive control group (wait-list, no-contact). In multi-arm studies, all eligible control conditions were included.

#### 2.3.5 Types of outcomes

Outcomes included were change from baseline to post-intervention on measures of untrained cognitive outcomes (global or domain-specific), performance-based physical exercise outcomes (aerobic capacity, strength, mobility, balance or gait), psychosocial outcomes (neuropsychiatric symptoms, depression, quality of life) and functional outcomes (activities of daily living (ADL) or instrumental ADL).

### 2.4 Data collection and coding

Data were extracted by one reviewer (HMG or CD) and checked against the original publication by another reviewer (SA). For each study, we extracted population, intervention and control characteristics and results for each eligible outcome. Outcomes were recorded as mean and standard deviation for each condition and time-point (pre and post intervention). When means and standard deviations were not available, data were entered as mean change and standard deviation or mean differences and 95% confidence interval (CI). If data could not be extracted from the study report, we contacted the authors and asked for missing group-level data. For data extraction purposes, cognitive outcomes were classified by a neuropsychologist (HMG) according to the Cattell-Horn-Carroll-Miyake taxonomy of cognitive domains described by Webb et al. (2018): (1) executive function; (2) short-term and working memory; (3) long-term storage and retrieval; (4) processing speed; (5) visual processing; and (6) fluid reasoning. Although this framework allows for further classification of cognitive outcomes into more specific narrow abilities (e.g., *short-term and working memory* can be further subdivided into *high working memory*, *low working memory* and *short-term memory*), we used the broad abilities to ensure a sufficient number of studies within each category to allow for meaningful interpretation. Cognitive screening instruments, such as the Mini-Mental State Examination, were classified as global cognition (Hill et al., 2017; Lampit et al., 2014). Physical outcomes were classified according to the following domains: (1) aerobic capacity; (2) strength; (3) functional mobility; (4) gait; (5) balance; and (6) cognitive-motor outcomes (concurrent physical and cognitive tasks), these classifications were made under the supervision of an exercise physiologist (AS). For cognitive-motor outcomes, only the physical performance outcome (i.e., not cognitive task performance) was included. The classifications of outcomes by domain are provided in Appendix B. Following the approach of previous meta-analyses (Gheysen et al., 2018; Stanmore et al., 2017; Zhu et al., 2016), each combined intervention arm was classified according to their mode of delivery as either simultaneous, sequential or exergaming training. Comparison groups were classified as: cognitive training (cognitive training alone or in combination with sham physical exercise, e.g., stretching), physical exercise (physical exercise alone or in combination with sham cognitive training or other non-specific cognitive activities), sham interventions (active control groups, e.g., sham physical exercise and/or sham cognitive training, health education, non-specific cognitive activities) or passive control (wait-list, no-contact). Study participants were classified as either cognitively healthy, MCI, dementia or Parkinson’s disease by examining the study inclusion criteria and the baseline characteristics of the sample. Studies including participants with subjective cognitive complaints but no objective cognitive impairment were classified as cognitively healthy. Classification of clinical populations were based on the definitions provided by the study authors; for MCI this could include diagnosis based on clinical assessment and/or cognitive cut-off scores. If a study included a cognitively or physically frail population without further specification of diagnosis (e.g., people in long-term nursing homes), we classified the population based on the cognitive and functional status at baseline (Hill et al., 2017).

### 2.5 Risk of bias within studies

Two independent reviewers (CD and RM) assessed the risk of bias in individual studies using the revised Cochrane risk-of-bias tool for randomized trials (ROB 2) (Sterne et al., 2019). Disagreements were resolved by consulting a third reviewer (HMG or AL). Studies with high risk of bias or some concerns in the domains bias due to missing outcome data or bias in measurement of the outcome were considered as having a high risk of bias or some concerns, respectively (Hill et al., 2017; Lampit et al., 2014).

### 2.6 Data analysis

Analyses were conducted using the R packages robumeta (Fisher et al., 2017), clubSandwich (Pustejovsky, 2020) and netmeta (Rücker et al., 2020a). Between-group differences in change from baseline to post-intervention were converted to standardized mean differences and calculated as Hedges’ *g* with 95% CI. Pooling of effect estimates across studies were conducted using random-effects models. Studies including participants with dementia and Parkinson’s disease were excluded from the pooled analyses due to an insufficient number of identified studies available for analysis (*k*=2 for dementia and Parkinson’s disease, respectively; see *3.2 Characteristics of included studies*). Instead, these studies were summarized narratively.

The analysis was conducted in two steps: First, pairwise meta-analysis was conducted using robust variance estimation implemented in robumeta, accounting for the dependency structure of effect estimates within studies (Hedges et al., 2010). Analyses were performed for overall cognitive (Gheysen et al., 2018; Hill et al., 2017; Lampit et al., 2014; Stanmore et al., 2017; Zhu et al., 2016), physical (Falck et al., 2019), psychosocial and functional outcomes (Hill et al., 2017) as well as for each cognitive and physical subdomain separately. Heterogeneity across studies was quantified using τ^2^ and expressed as a proportion of overall observed variance using the I^2^ statistic (Borenstein et al., 2017). Prediction intervals were calculated to assess the dispersion of true effects (Riley et al., 2011). Subgroup analyses based on pre-specified categorical moderators (population group, intervention duration, supervision, risk of bias) were conducted using mixed-effects meta-regression models. Small-study effect was investigated by visually inspecting funnel plots of effect size vs standard error and formally tested using the Egger’s test (Egger et al., 1997; Sterne et al., 2011).

Second, random-effects network meta-analysis was performed using a frequentist framework, implemented in the netmeta package for R. To examine the transitivity assumptions, we created a table summarizing potential effect modifiers (population and design characteristics) to explore whether these were similarly distributed across the different comparisons. The geometry of the network was summarized in a network graph and league tables were created to display the relative effect sizes of all available comparisons. Ranking of treatments were estimated using P-scores, representing the extent of certainty that an intervention is more effective than another intervention, averaged over all treatment arms (Rucker and Schwarzer, 2015). Sensitivity analyses were conducted excluding trials with a high risk of bias, and by conducting separate analyses for studies classified as cognitively healthy older adults and MCI. The certainty of the evidence was investigated using the Confidence in Network Meta-Analysis tool (Nikolakopoulou et al., 2020; Papakonstantinou et al., 2020), incorporating six domains: within-study bias, reporting bias, indirectness, imprecision, heterogeneity and incoherence.

## 3. Results

### 3.1 Study selection

After exclusion of duplicate search results, we screened 23,102 articles for eligibility, of which 22,690 were excluded based on titles and abstracts. Consequently, 412 articles were assessed in full-text screening. Of these, 45 unique studies fulfilled inclusion criteria. A list of the excluded studies (with reasons) is provided in Appendix C. Two additional eligible studies were identified through manual search (Adcock et al., 2020; Rezola-Pardo et al., 2019). Thus, a total of 47 studies fulfilled inclusion criteria (Fig. 1). Additionally, eight secondary outcome articles were identified (Boa Sorte Silva et al., 2018a; Boa Sorte Silva et al., 2017; Eggenberger et al., 2015b; Fraser et al., 2017; Hagovska and Nagyova, 2017; Hagovska and Olekszyova, 2016; Mavros et al., 2017; Middleton et al., 2018) and included along with other manuscripts from the same study. The authors of seven studies (Barban et al., 2017; Boa Sorte Silva et al., 2018b; Htut et al., 2018; Mavros et al., 2017; McDaniel et al., 2014; Middleton et al., 2018; Ten Brinke et al., 2019) were contacted for data and four (Htut et al., 2018; Mavros et al., 2017; Middleton et al., 2018; Ten Brinke et al., 2019) provided data.

**Fig. 1.**
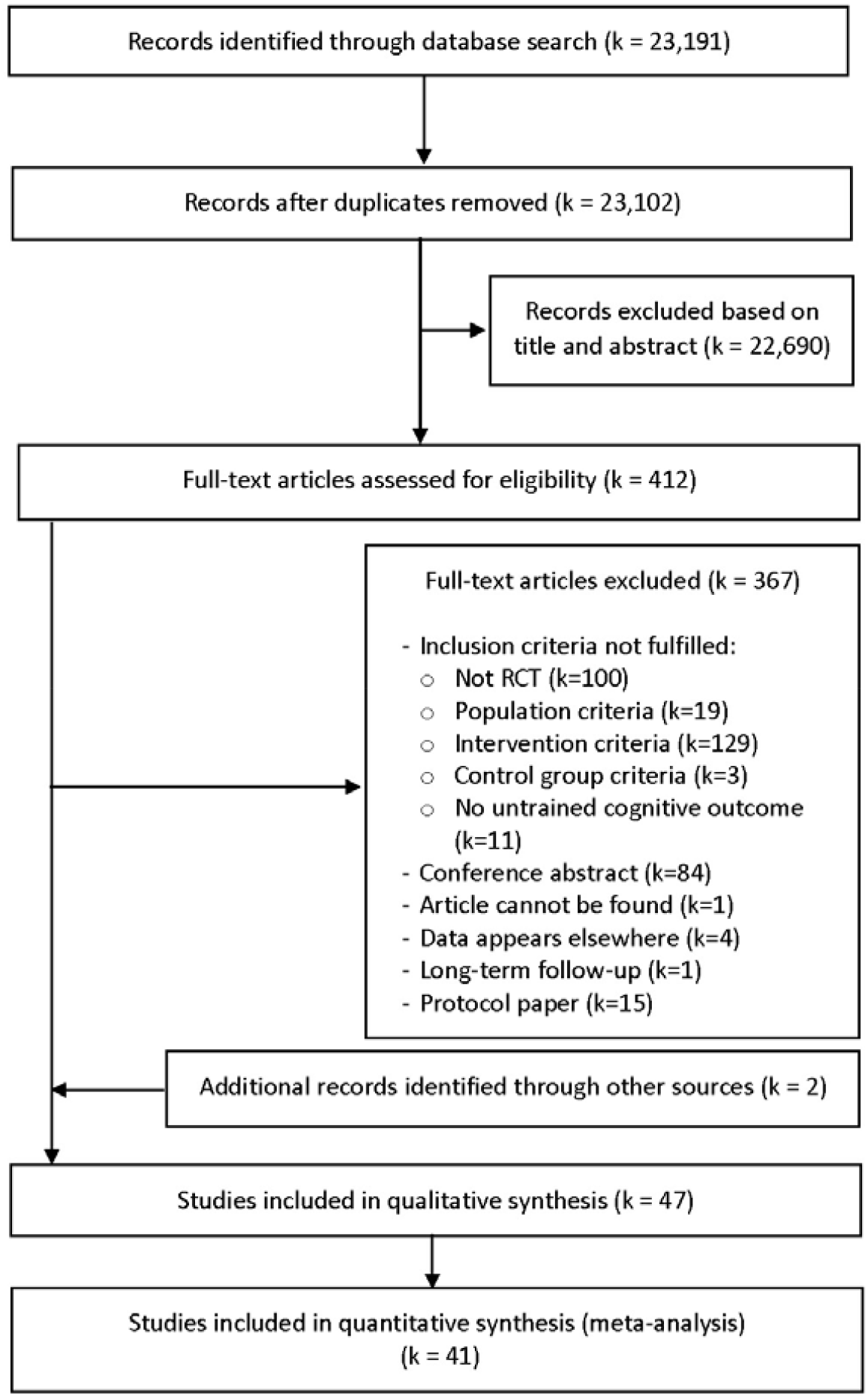
PRISMA flow chart

### 3.2 Characteristics of included studies

The characteristics of the included studies are presented in Table 1. The included studies encompassed 4,052 participants with mean age ranging between 65.0 and 87.2 years. The majority of the studies included cognitively healthy older adults (*k*=30, *n*=2,866), two of which (Barnes et al., 2013; Boa Sorte Silva et al., 2018b) included participants with subjective cognitive complaints. Twelve studies included participants with MCI and one additional study (Stanmore et al., 2019) included people living in assisted living facilities without severe cognitive impairment. This study was classified as MCI for the purpose of the subgroup analyses, giving a total of 13 studies (*n*=932) in this category. Two studies included participants with Parkinson’s disease without dementia (Pompeu et al., 2012; Song et al., 2018). One study included people with dementia (Karssemeijer et al., 2019) and one included long-term nursing home residents (Rezola-Pardo et al., 2019), which was classified as dementia and thus excluded from the pooled analyses. Intervention duration ranged from four to 40 weeks (median = 12 weeks). Sixteen studies combined cognitive and physical training using a sequential design, in which eight provided physical and cognitive training in separate sessions (Barnes et al., 2013; Desjardins-Crepeau et al., 2016; Fabre et al., 2002; Maffei et al., 2017; McDaniel et al., 2014; Romera-Liebana et al., 2018; Shatil, 2013; van het Reve and de Bruin, 2014), and eight implemented the cognitive and physical training back-to-back within the same session (Barban et al., 2017; Damirchi et al., 2018; Fiatarone Singh et al., 2014; Hagovska et al., 2016; Legault et al., 2011; Linde and Alfermann, 2014; Rahe et al., 2015; Ten Brinke et al., 2019). Examples of sequential training include separate sessions of aerobic and resistance training, and computerized cognitive training (see Table 1 for details). Thirteen studies used a simultaneous design (Boa Sorte Silva et al., 2018b; Combourieu Donnezan et al., 2018; Hiyamizu et al., 2012; Kitazawa et al., 2015; Laatar et al., 2018; Leon et al., 2015; Mrakic-Sposta et al., 2018; Nishiguchi et al., 2015; Norouzi et al., 2019; Park et al., 2019; Reigal and Mendo, 2014; Rezola-Pardo et al., 2019; Shimada et al., 2018), such as learning complex stepping patterns, solving cognitive tasks while simultaneously performing strength and balance exercises, or conducting computerized cognitive training during aerobic exercise on bikes. Seventeen studies included exergaming interventions (Adcock et al., 2020; Anderson-Hanley et al., 2018; Bacha et al., 2018; Barcelos et al., 2015; Delbroek et al., 2017; Eggenberger et al., 2016; Gschwind et al., 2015; Htut et al., 2018; Hughes et al., 2014; Karssemeijer et al., 2019; Maillot et al., 2012; Pompeu et al., 2012; Schattin et al., 2016; Schoene et al., 2013; Schoene et al., 2015; Song et al., 2018; Stanmore et al., 2019), examples include videogame dancing, cybercycling exergames, and commercial videogames with cognitively challenging components. One study (Eggenberger et al., 2015a) included an exergaming as well as a simultaneous intervention, while another (Rahe et al., 2015) included two sequential training intervention arms, one of which also included motivational counselling. The latter study was split into two separate studies for the network meta-analysis to allow for inclusion of both sequential training arms in the analysis. Comparison conditions included physical exercise (*k*=29), cognitive training (*k*=13), sham interventions (*k*=14) and passive control (*k*=19). Ten studies (21%) had a high risk of bias, 17 (36%) had some concerns, and 20 (43%) had a low risk of bias (Appendix D).

**Table 1.**
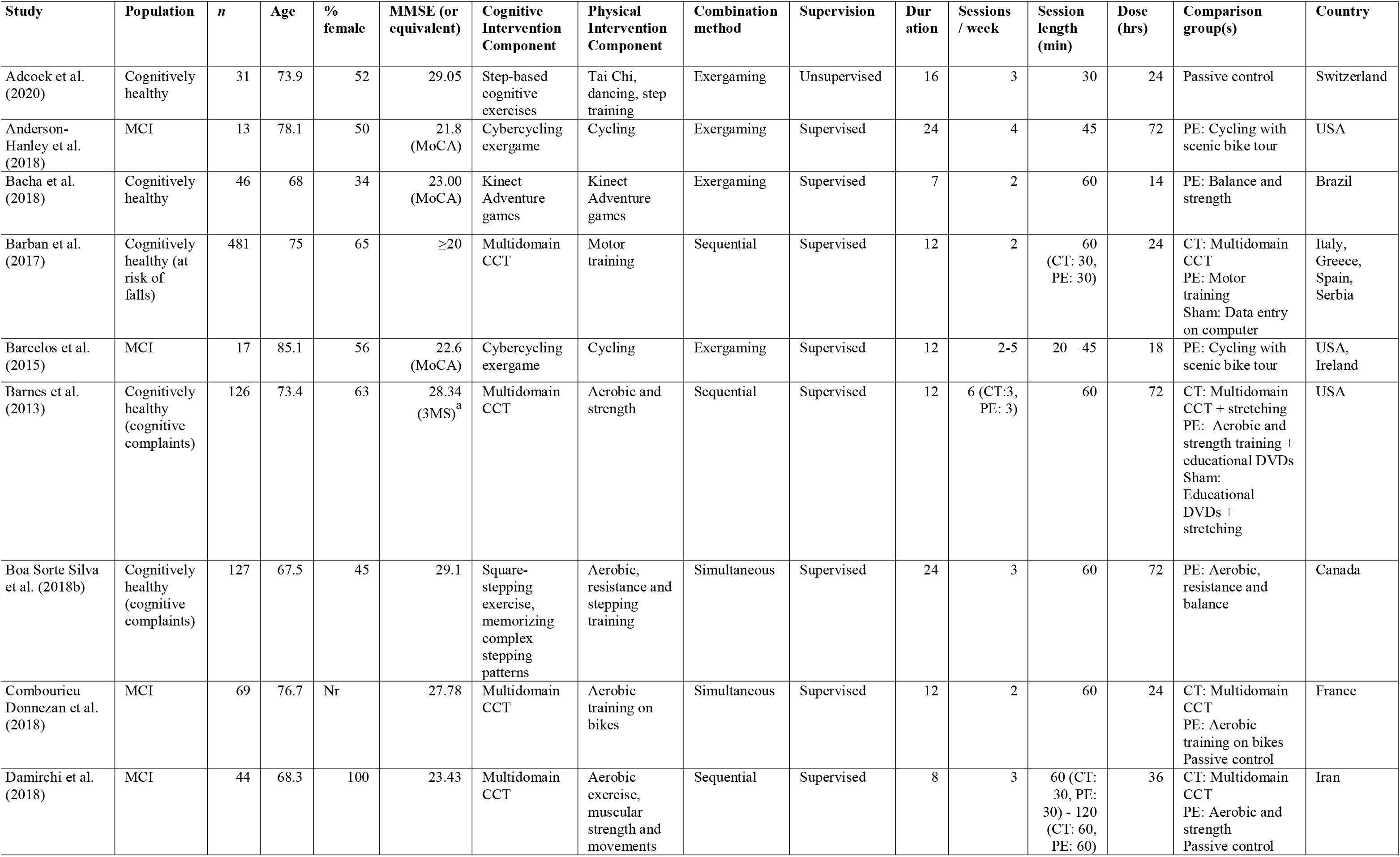
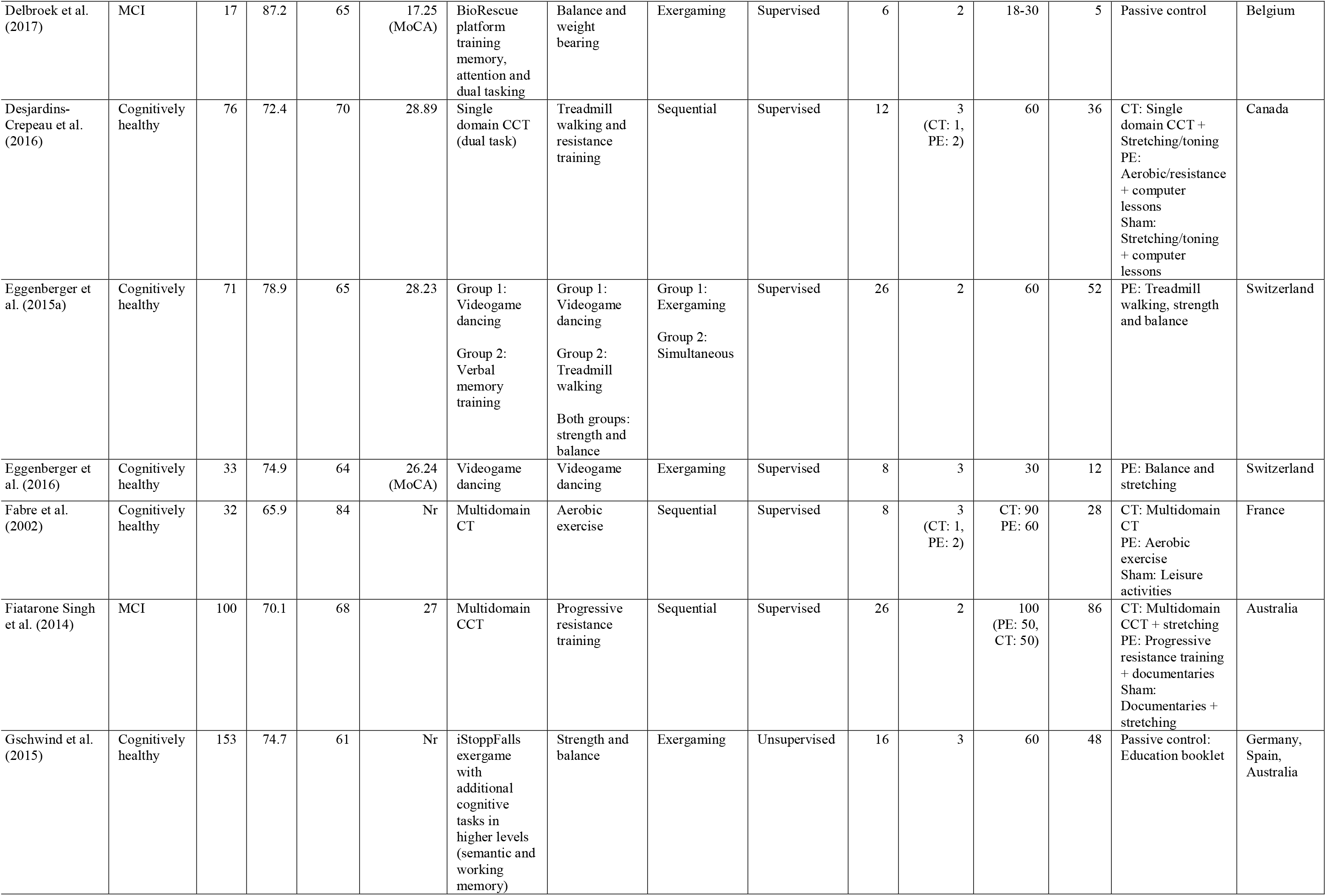
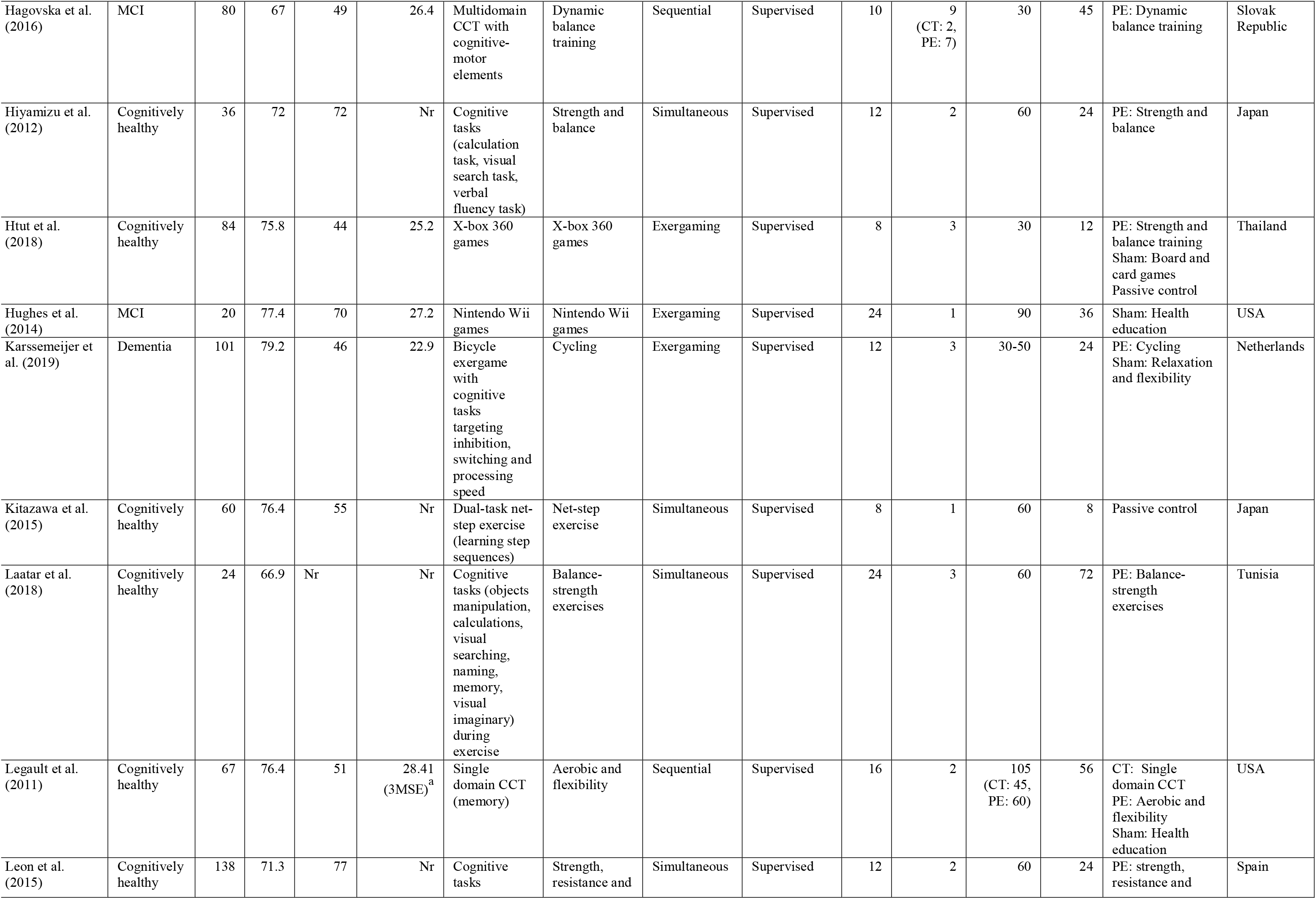
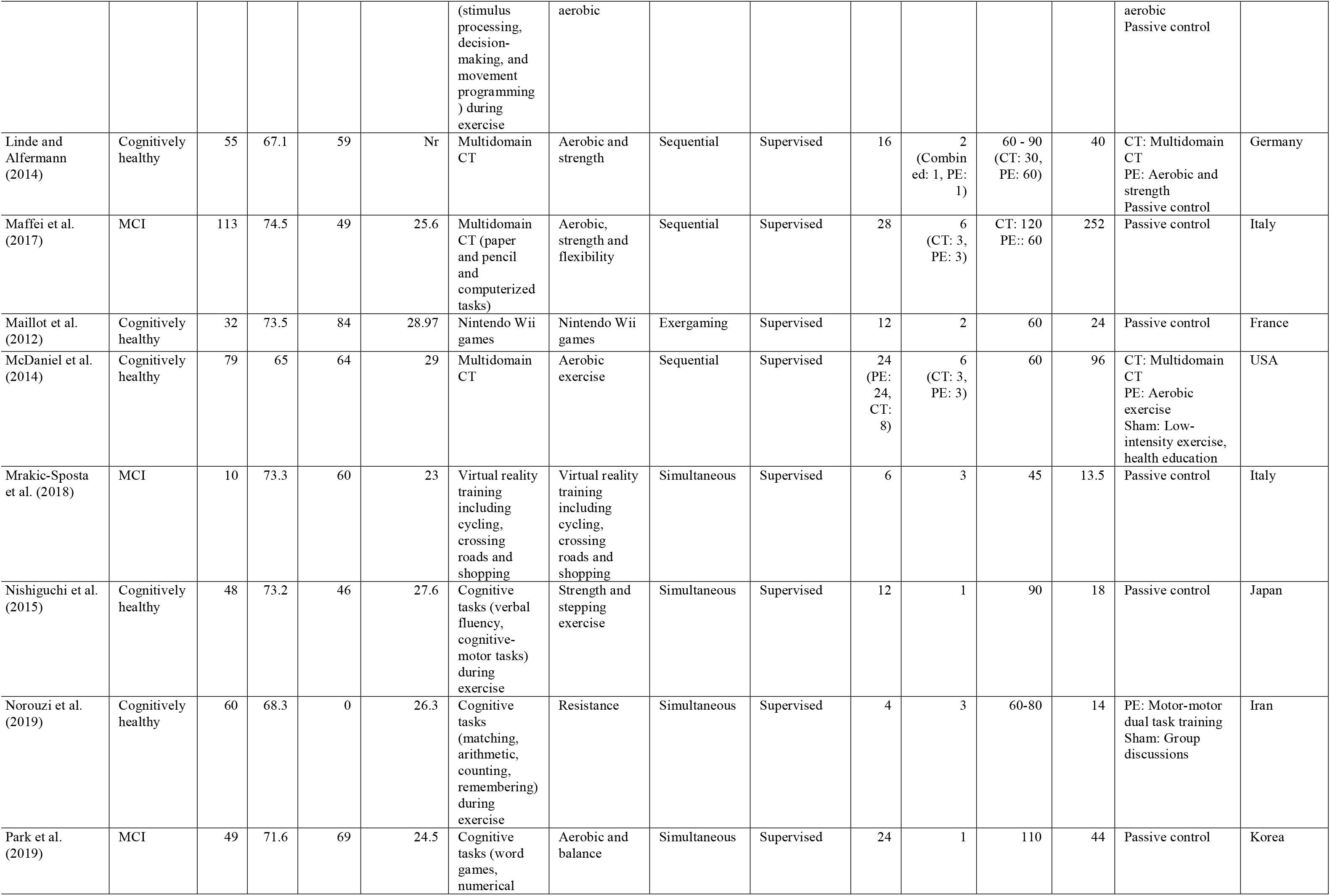
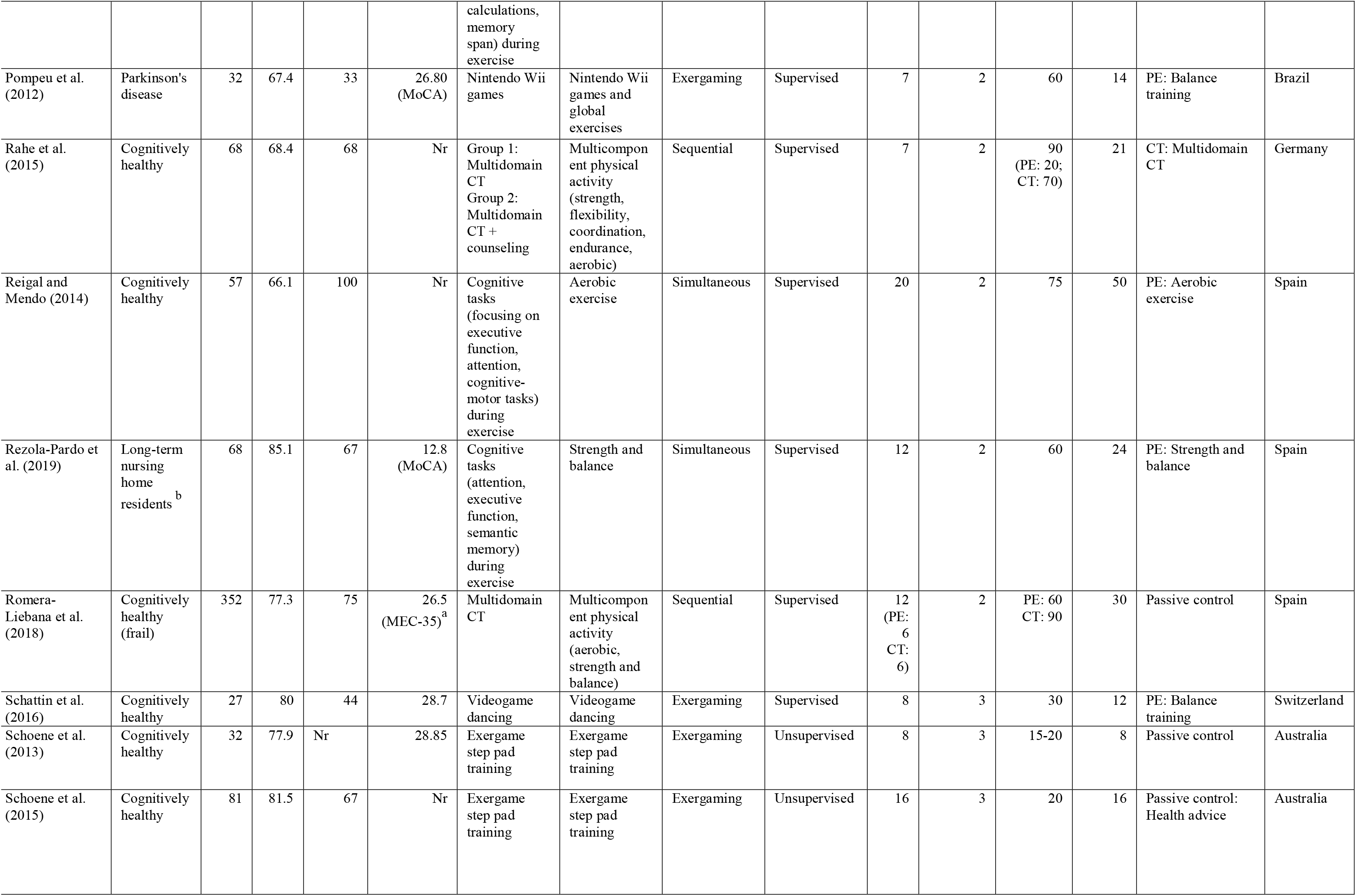
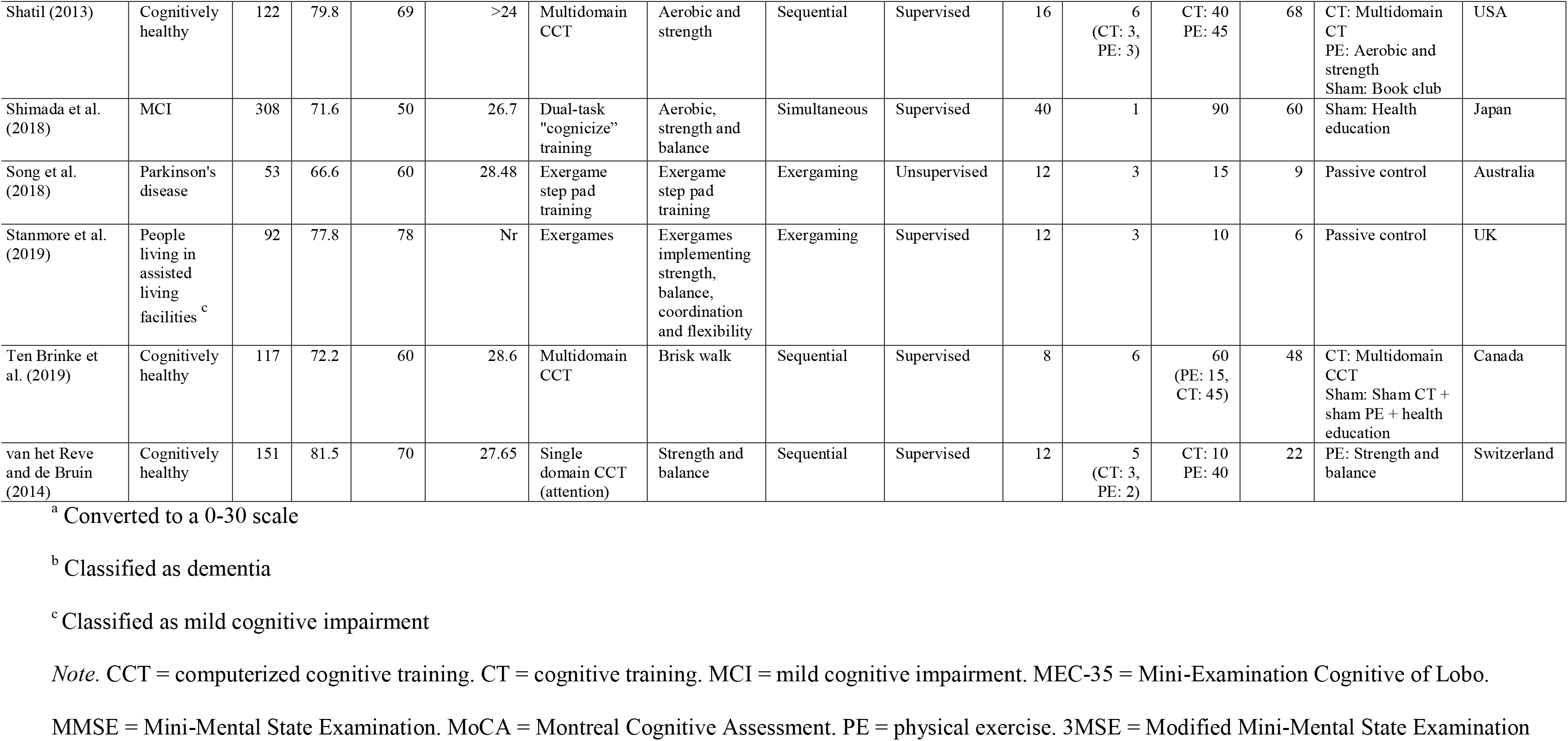
Characteristics of Included Studies

### 3.4 Pairwise meta-analysis

#### 3.4.1 Cognitive function

Across 43 studies and 491 effect estimates, the overall effect of combined interventions on cognitive function was moderate and statistically significant, Hedges’ *g* = 0.34 (95% CI 0.14 to 0.55), with high heterogeneity (τ^2^ = 0.29, I^2^ = 82%). The funnel plot revealed two conspicuous outliers, both reporting implausibly large *g* values of 1.74 (Laatar et al., 2018) and 4.52-5.54 (Leon et al., 2015) under high risk of bias. The studies were therefore removed from all further analysis. After removal of these outliers, heterogeneity was reduced, and the funnel plot showed no significant asymmetry (β = 0.51, *p* = 0.24, Appendix E). Across the remaining 41 studies and 482 effect estimates, the overall effect on cognitive outcomes was small and statistically significant, *g* = 0.22 (95% CI 0.14 to 0.30, τ^2^ = 0.08, I^2^ = 56%, prediction interval −0.34 to 0.78). The results of individual studies and comparisons are provided in Appendix F. There was no statistically significant difference in effect estimates between studies classified as low, some concerns or high risk of bias (Table 2).

**Table 2.**
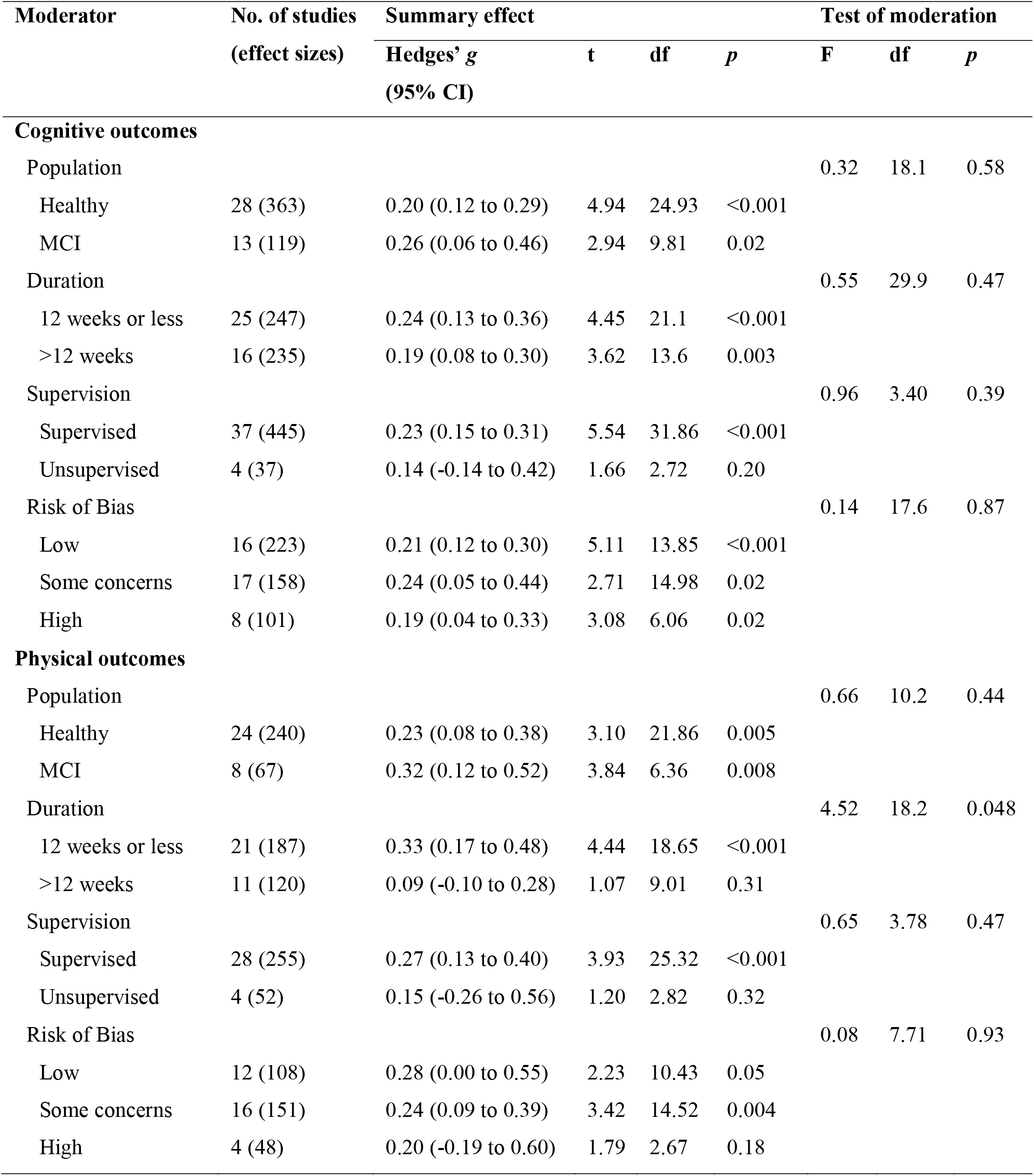
Moderator analyses of overall cognitive and physical outcomes.

| Moderator | No. of studies<br>(effect sizes) | Summary effect |  |  |  | Test of moderation |  |  |
| --- | --- | --- | --- | --- | --- | --- | --- | --- |
|  |  | Hedges' *g*<br>(95% CI) | *t* | *df* | *p* | *F* | *df* | *p* |
| Cognitive outcomes |  |  |  |  |  |  |  |  |
| Population |  |  |  |  |  | 0.32 | 18.1 | 0.58 |
| Healthy | 28 (363) | 0.20 (0.12 to 0.29) | 4.94 | 24.93 | <0.001 |  |  |  |
| MCI | 13 (119) | 0.26 (0.06 to 0.46) | 2.94 | 9.81 | 0.02 |  |  |  |
| Duration |  |  |  |  |  | 0.55 | 29.9 | 0.47 |
| 12 weeks or less | 25 (247) | 0.24 (0.13 to 0.36) | 4.45 | 21.1 | <0.001 |  |  |  |
| >12 weeks | 16 (235) | 0.19 (0.08 to 0.30) | 3.62 | 13.6 | 0.003 |  |  |  |
| Supervision |  |  |  |  |  | 0.96 | 3.40 | 0.39 |
| Supervised | 37 (445) | 0.23 (0.15 to 0.31) | 5.54 | 31.86 | <0.001 |  |  |  |
| Unsupervised | 4 (37) | 0.14 (-0.14 to 0.42) | 1.66 | 2.72 | 0.20 |  |  |  |
| Risk of Bias |  |  |  |  |  | 0.14 | 17.6 | 0.87 |
| Low | 16 (223) | 0.21 (0.12 to 0.30) | 5.11 | 13.85 | <0.001 |  |  |  |
| Some concerns | 17 (158) | 0.24 (0.05 to 0.44) | 2.71 | 14.98 | 0.02 |  |  |  |
| High | 8 (101) | 0.19 (0.04 to 0.33) | 3.08 | 6.06 | 0.02 |  |  |  |
| Physical outcomes |  |  |  |  |  |  |  |  |
| Population |  |  |  |  |  | 0.66 | 10.2 | 0.44 |
| Healthy | 24 (240) | 0.23 (0.08 to 0.38) | 3.10 | 21.86 | 0.005 |  |  |  |
| MCI | 8 (67) | 0.32 (0.12 to 0.52) | 3.84 | 6.36 | 0.008 |  |  |  |
| Duration |  |  |  |  |  | 4.52 | 18.2 | 0.048 |
| 12 weeks or less | 21 (187) | 0.33 (0.17 to 0.48) | 4.44 | 18.65 | <0.001 |  |  |  |
| >12 weeks | 11 (120) | 0.09 (-0.10 to 0.28) | 1.07 | 9.01 | 0.31 |  |  |  |
| Supervision |  |  |  |  |  | 0.65 | 3.78 | 0.47 |
| Supervised | 28 (255) | 0.27 (0.13 to 0.40) | 3.93 | 25.32 | <0.001 |  |  |  |
| Unsupervised | 4 (52) | 0.15 (-0.26 to 0.56) | 1.20 | 2.82 | 0.32 |  |  |  |
| Risk of Bias |  |  |  |  |  | 0.08 | 7.71 | 0.93 |
| Low | 12 (108) | 0.28 (0.00 to 0.55) | 2.23 | 10.43 | 0.05 |  |  |  |
| Some concerns | 16 (151) | 0.24 (0.09 to 0.39) | 3.42 | 14.52 | 0.004 |  |  |  |
| High | 4 (48) | 0.20 (-0.19 to 0.60) | 1.79 | 2.67 | 0.18 |  |  |  |

The results of the moderator analyses for cognitive outcomes are shown in Table 2. A statistically significant effect for was found for cognitively healthy older adults and for participants with MCI, with no significant difference between the groups. There was no significant difference in the efficacy of interventions that were 12 weeks or shorter compared to interventions with a duration of > 12 weeks. Similarly, no evidence for statistically significant subgroup difference was found for supervised training compared to unsupervised training, however, because of the small number of studies with unsupervised training (i.e., df<4), these results should be interpreted with caution. For the different cognitive domains, statistically significant effect estimates were found for executive function, short-term and working memory, long-term storage and retrieval, processing speed, fluid reasoning and global cognition, but not for visual processing (Table 3).

**Table 3.** Meta-analyses of cognitive and physical domains.

| Domain | No. of studies<br>(effect sizes) | Hedges' $g$ (95% CI) | $I^2$ | $\tau^2$ |
| --- | --- | --- | --- | --- |
| **Cognitive outcomes** |  |  |  |  |
| Executive function | 27 (113) | 0.22 (0.13 to 0.30) | 31% | 0.03 |
| Short-term and working memory | 21 (75) | 0.30 (0.05 to 0.55) | 72% | 0.18 |
| Long-term storage and retrieval | 18 (119) | 0.17 (0.04 to 0.30) | 47% | 0.04 |
| Processing speed | 22 (109) | 0.17 (0.07 to 0.27) | 30% | 0.03 |
| Visual processing | 6 (14) | 0.11 (-0.18 to 0.40) | 59% | 0.12 |
| Fluid reasoning | 7 (25) | 0.24 (0.02 to 0.46) | 36% | 0.03 |
| Global cognition | 15 (22) | 0.30 (0.06 to 0.53) | 70% | 0.15 |
| **Physical outcomes** |  |  |  |  |
| Functional mobility | 22 (83) | 0.34 (0.16 to 0.52) | 70% | 0.15 |
| Strength | 9 (33) | 0.17 (-0.10 to 0.44) | 55% | 0.07 |
| Aerobic capacity | 14 (39) | 0.10 (-0.06 to 0.26) | 13% | 0.01 |
| Balance | 12 (24) | 0.23 (-0.04 to 0.49) | 62% | 0.09 |
| Gait | 10 (41) | 0.06 (-0.23 to 0.34) | 60% | 0.12 |
| Cognitive-motor | 14 (87) | 0.19 (-0.01 to 0.39) | 64% | 0.12 |

#### 3.4.2 Physical function

Thirty-two studies reporting 307 effect estimates were available for physical outcomes. The overall effect was small and statistically significant, *g* = 0.25 (95% CI 0.13 to 0.37), with moderate heterogeneity (τ^2^ = 0.12, I^2^ = 63%, 95% prediction interval −0.46 to 0.96). The results of individual studies and comparisons are provided in Appendix G. There was no indication of funnel plot asymmetry (β= 0.47, *p* = 0.52, Appendix E). No statistically significant difference was found in effect estimates between studies classified as low, some concerns or high risk of bias (Table 2).

There was no statistically significant difference in the efficacy of combined interventions on physical outcomes for cognitively healthy older adults compared to those with MCI. A significant moderating effect was found for intervention duration, whereby a beneficial effect was seen for interventions that were 12 weeks or shorter but not for interventions with > 12 weeks duration. No significant moderating effect on physical outcomes was seen for training supervision; however, due to the small number of studies with unsupervised training, these results should be interpreted with caution. Table 2 shows the full results from the moderator analyses of physical outcomes. For the physical subdomains, a statistically significant effect was found for functional mobility, but not for strength, aerobic capacity, balance, gait, or cognitive-motor outcomes (Table 3).

#### 3.4.3 Psychosocial function

Nine studies reporting 20 psychosocial effect estimates were available, six in cognitively healthy older adults and three in MCI. The results of individual studies and comparisons are provided in Appendix H. The overall effect was small and statistically non-significant, *g* = 0.28 (95% CI −0.16 to 0.72) with high heterogeneity (τ^2^ = 0.24, I^2^ = 81%, prediction interval −0.97 to 1.53). Visual inspection of the funnel plot revealed one outlier (Hagovska et al., 2016) (see Appendix E), reporting an effect estimate of *g* = 1.80. A sensitivity analysis after removal of the outlier revealed a negligible but statistically significant effect, *g* = 0.10 (95% CI 0.01 to 0.20) with no heterogeneity (τ^2^ = 0.00, I^2^ = 0%).

#### 3.4.4 Functional abilities

Only two trials were identified that reported functional outcomes (Fiatarone Singh et al., 2014; Hagovska et al., 2016) and data were therefore not pooled in meta-analysis. Both studies included participants with MCI. Improvements following combined interventions were reported for ADL (Hagovska and Nagyova, 2017) but not for instrumental ADL (Fiatarone Singh et al., 2014; Hagovska et al., 2016). One additional study (Gschwind et al., 2015) reported no significant benefit in general health and disability in healthy older adults.

#### 3.4.5 Studies in dementia or Parkinson’s disease

The results for dementia (*k*=2) and Parkinson’s disease (*k*=2) are provided in Appendix I, showing no significant benefits on overall cognitive, physical or psychosocial function in the individual studies and comparisons. For dementia, Karssemeijer et al. (2019) reported improvements in psychomotor speed following an exergaming intervention compared to active control, but not compared to physical exercise alone. No differences between the groups were found in working memory, executive function or episodic memory. Rezola-Pardo et al. (2019) reported no additional benefits of simultaneous training compared to physical exercise alone on a range of cognitive, physical and psychosocial outcomes in people living in long-term nursing homes. In Parkinson’s disease, two studies reported no cognitive or physical benefits of exergaming compared to physical exercise (Pompeu et al., 2012) or passive control (Song et al., 2018).

### 3.5 Network meta-analysis

One study (Htut et al., 2018) was excluded from the network meta-analysis due to inconsistent treatment estimates and variances. Visual inspection of potential effect modifiers showed that these were similarly distributed across the network of included comparisons (Appendix J), suggesting that the transitivity assumption was plausible. Four studies used unsupervised training, all of which compared exergaming with passive control (Adcock et al., 2020; Gschwind et al., 2015; Schoene et al., 2013; Schoene et al., 2015). These studies were therefore downgraded to moderate indirectness in the grading of the certainty of the evidence. Figure 2 visualizes the well-connected network structure for both cognitive (Fig. 2a) and physical (Fig. 2b) outcomes, indicating a high amount of direct evidence for the various treatment comparisons. The most frequently examined comparisons were between sequential training, physical exercise, cognitive training and sham intervention, as well as between exergaming and passive control.

**Fig. 2.**
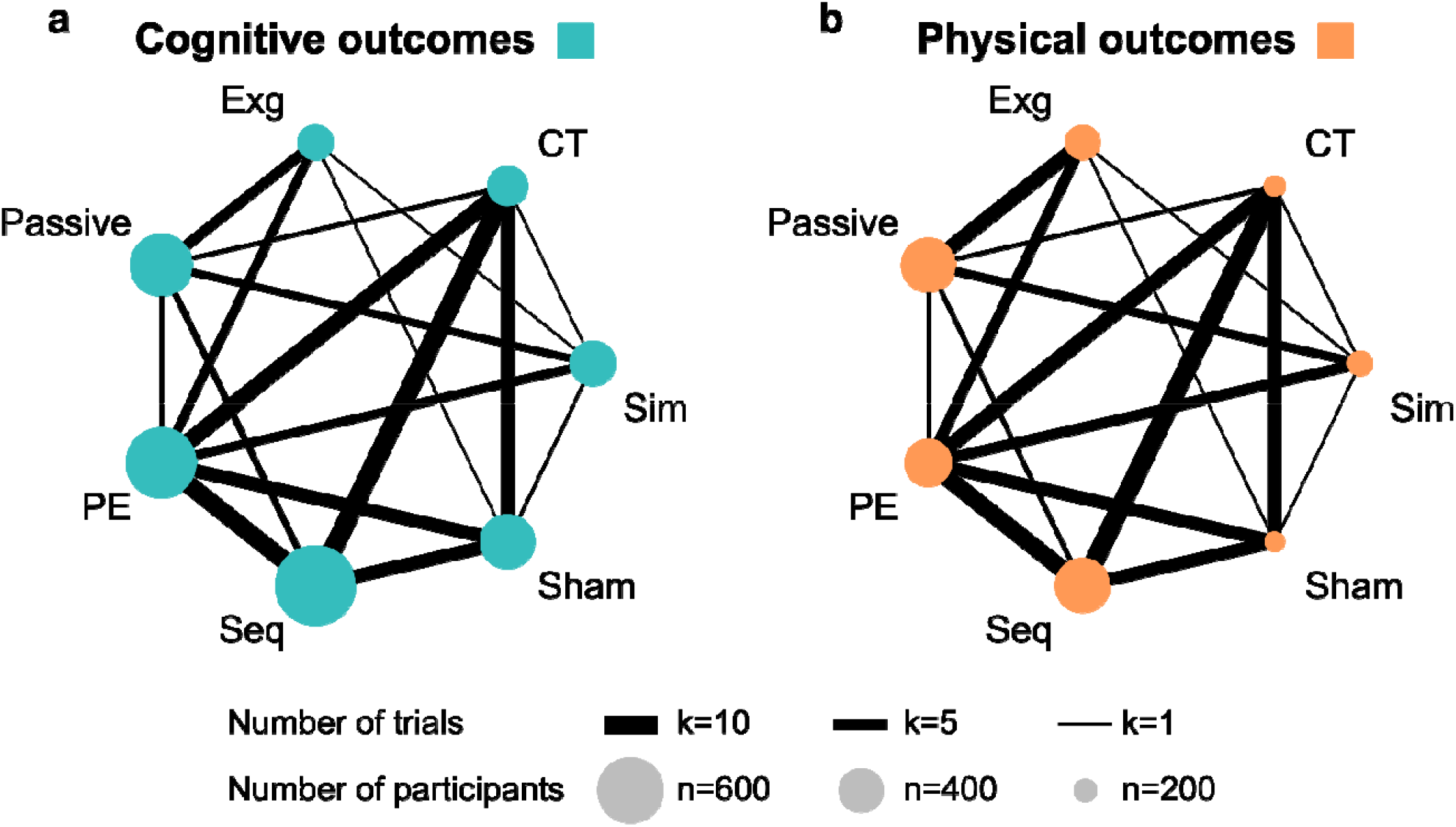
Network plot for (a) cognitive outcomes and (b) physical outcomes. The width of the lines represents the number of studies comparing each pair of treatments. The size of the circle represents the sample size in each arm. CT = cognitive training. Exg = exergaming. Passive = passive control. PE = physical exercise. Seq = sequential training. Sham = sham intervention. Sim = simultaneous training.

#### 3.5.1 Cognitive function

Network meta-analysis for cognitive outcomes included 41 studies, 28 in cognitively healthy older adults and 13 in MCI. The results are shown in Table 4. Indirect evidence was obtained for four comparisons for which direct evidence was unavailable (exergaming vs cognitive training, exergaming vs sequential training, sham intervention vs passive control, and sequential vs simultaneous training, Fig 2a). All conditions except sham interventions were significantly more efficacious for cognitive function than passive control (*g* range 0.18 to 0.43). Simultaneous and sequential training were significantly more efficacious for cognitive function than physical exercise (*g* = 0.24 and 0.15, respectively) and sham intervention (*g* = 0.31 and 0.22, respectively). A statistically significant benefit was also found for simultaneous training relative to exergaming (*g* = 0.25). No significant differences were found between the three types of combined interventions compared to cognitive training alone (*g* range −0.14 to 0.11). The certainty of the evidence is shown in Appendix K. Confidence ratings were moderate or high for the majority of the comparisons, with the exception of low certainty of the evidence due to within study bias and imprecision for the comparisons of exergaming vs sequential training, cognitive training and sham interventions; and simultaneous training vs sequential training and cognitive training.

**Table 4.** Relative effect estimates for the contrasts between the different intervention and control arms on cognitive function (below diagonal) and physical function (above diagonal). Statistically significant effects are shown in bold.

|  |  |  |  |  |  |  |
| --- | --- | --- | --- | --- | --- | --- |
| **Simultaneous** | 0.01<br>[-0.22; 0.24] | 0.23<br>[-0.01; 0.47] | **0.33**<br>**[0.07; 0.59]** | 0.12<br>[-0.08; 0.32] | **0.35**<br>**[0.08; 0.62]** | **0.44**<br>**[0.23; 0.66]** |
| 0.09<br>[-0.06; 0.25] | **Sequential** | 0.22<br>[-0.004; 0.44] | **0.32**<br>**[0.13; 0.51]** | 0.11<br>[-0.05; 0.27] | **0.34**<br>**[0.13; 0.55]** | **0.43**<br>**[0.24; 0.63]** |
| **0.25**<br>**[0.06; 0.45]** | 0.16<br>[-0.02; 0.33] | **Exergaming** | 0.10<br>[-0.16; 0.36] | -0.11<br>[-0.32; 0.09] | 0.12<br>[-0.15; 0.39] | **0.21**<br>**[0.04; 0.38]** |
| 0.11<br>[-0.06; 0.28] | 0.02<br>[-0.11; 0.14] | -0.14<br>[-0.34; 0.06] | **Cognitive training** | **-0.21**<br>**[-0.41; -0.01]** | 0.02<br>[-0.20; 0.24] | 0.11<br>[-0.13; 0.36] |
| **0.24**<br>**[0.10; 0.38]** | **0.15**<br>**[0.03; 0.26]** | -0.01<br>[-0.18; 0.16] | 0.13<br>[-0.003; 0.26] | **Physical exercise** | **0.23**<br>**[0.02; 0.44]** | **0.32**<br>**[0.13; 0.52]** |
| **0.31**<br>**[0.17; 0.46]** | **0.22**<br>**[0.09; 0.35]** | 0.06<br>[-0.13; 0.25] | **0.20**<br>**[0.07; 0.34]** | 0.07<br>[-0.05; 0.20] | **Sham** | 0.09<br>[-0.16; 0.35] |
| **0.43**<br>**[0.27; 0.60]** | **0.34**<br>**[0.21; 0.47]** | **0.18**<br>**[0.03; 0.33]** | **0.32**<br>**[0.16; 0.49]** | **0.19**<br>**[0.05; 0.34]** | 0.12<br>[-0.04; 0.28] | **Passive** |

#### 3.5.2 Physical function

The network meta-analysis for physical outcomes included 32 studies, 24 in cognitively healthy older adults and eight in MCI. Indirect evidence was calculated for the same four comparisons as above for which direct evidence was unavailable. Simultaneous training, sequential training, exergaming and physical exercise were all significantly more efficacious for physical function than passive control (*g* range 0.21 to 0.44, Table 2). Simultaneous training, sequential training and physical exercise were significantly more efficacious for physical function than cognitive training (*g* range 0.21 to 0.33) and sham intervention (*g* range 0.23 to 0.35). Certainty of the evidence was low or very low for the majority of the comparisons, mainly due to within-study bias, imprecision and heterogeneity, with the exception of high certainty for the comparison between sequential training and passive control and moderate certainty for the comparison between simultaneous training and passive control; and physical exercise and passive control (Appendix K).

#### 3.5.3 Ranking of interventions

Figure 3 shows the relative rankings of the interventions with passive control as the reference treatment. For cognitive function, simultaneous training ranked best (P-score 0.96, Hedges’ *g* 0.43) followed by sequential training (P-score 0.78, Hedges’ *g* 0.34) and cognitive training (P-score 0.73, Hedges’ *g* 0.32). For physical function, simultaneous training ranked best (P-score 0.90, Hedges’ *g* 0.44), followed by sequential training (P-score 0.89, Hedges’ *g* 0.43) and physical exercise (P-score 0.67, Hedges’ g 0.32).

**Fig. 3.**
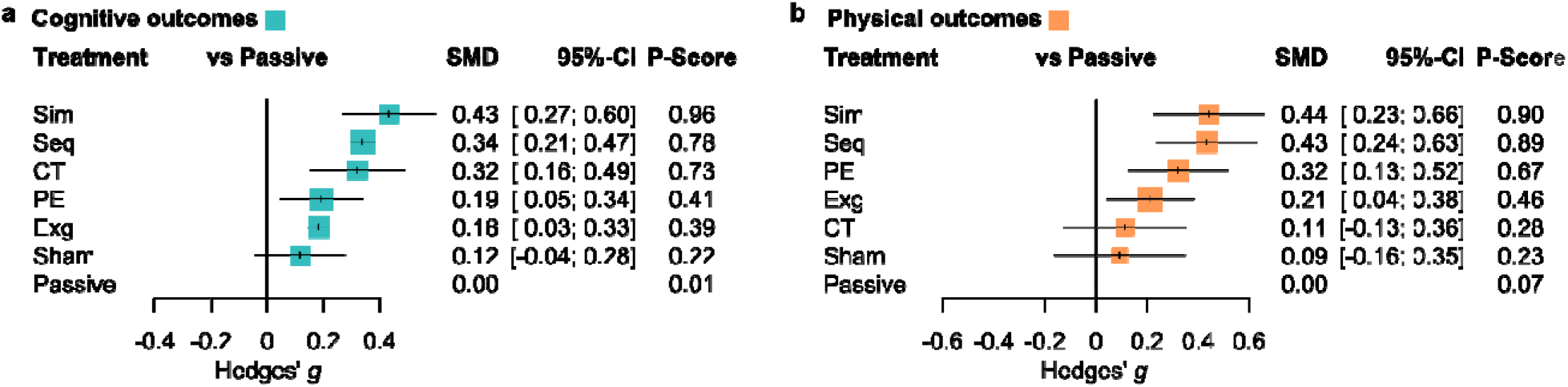
Forest plot showing the relative ranking and effect estimate of interventions with passive control as reference for overall (a) cognitive and (b) physical outcomes. CT = cognitive training. Exg = exergaming. Passive = passive control. PE = physical exercise. Seq = sequential training. Sham = sham intervention. Sim = simultaneous training.

#### 3.5.4 Sensitivity analyses

The results from the sensitivity analyses are shown in Appendix L. The first sensitivity analysis excluded studies with a high risk of bias. In the second sensitivity analysis, we performed separate analyses for studies with cognitively healthy older adults and MCI. Overall, results were highly convergent to the main analysis, although the small number of studies including participants with MCI (*k*=13 for cognitive outcomes and *k*=8 for physical outcomes) reduced the precision of effect estimates.

## 4. Discussion

Across 41 RCTs of narrowly defined combined cognitive training and physical exercise interventions, we found that in cognitively healthy older adults and those with MCI, simultaneous and sequential training lead to comparable cognitive benefits as cognitive training alone, while their efficacy on physical function is similar to that of physical exercise. Thus, although combined physical and cognitive interventions do not seem to induce additive cognitive effects beyond that of cognitive training, their physical efficacy make these a compelling strategy for maintaining cognitive alongside physical health in late life. Given the average dose of simultaneous training was substantially lower than that of sequential designs (mean total training hours: 33 [range 8 to 72] and 60 [range 21 to 252], respectively) with comparable efficacy, the former might be considered as a more feasible option in clinical and community settings. Conversely, our results reveal small effect estimates and lower relative rankings for exergaming and thus do not support the efficacy of such interventions, at least at this stage.

### 4.1 Interpretation of results and comparison with previous research

Our results provide a significant update to the current understanding of the effects of combined cognitive and physical training. We confirm the positive effects of combined interventions on cognition reported in previous meta-analyses (Gheysen et al., 2018; Stanmore et al., 2017; Zhu et al., 2016) and extend these findings by showing benefits also for overall physical function. Cognitive gains were found across most cognitive domains, whereas for physical function, a statistically significant effect was only observed for functional mobility, which was also the most frequently investigated outcome category. The overall cognitive and physical benefits were similar for cognitively healthy older adults and those with MCI and were not substantially influenced by bias within or across studies. Cognitive effect estimates did not depend on training duration; however, for physical outcomes, significant benefits were only observed for shorter (≤12 weeks) interventions. No evidence for a moderating effect of training supervision was found, although this should be interpreted with caution given that the majority of the included studies involved supervised training. Moreover, moderate heterogeneity was observed in several of the domain-specific pairwise meta-analyses; therefore, a more detailed investigation of potential effect modifiers, such as training content, dose, frequency and intensity, might be warranted to outline the intervention and design characteristics that could benefit specific cognitive and physical abilities.

Conversely, we did not find robust evidence that combined interventions improve psychosocial function, nor did we identify enough studies reporting on functional outcomes to synthesize the results in a pooled analysis. Although a statistically significant effect was observed for psychosocial function after outlier removal, the effect size was small and likely not of clinical relevance. Previous systematic reviews have reported mixed results for psychosocial and functional outcomes (Bruderer-Hofstetter et al., 2018; Karssemeijer et al., 2017; Zhu et al., 2016). The efficacy of combined interventions on everyday function and well-being therefore warrants further investigation and likely requires more consistent inclusion of these outcomes in primary trials in order to examine the extent to which these interventions can improve a wider range of clinically meaningful endpoints (Gavelin et al., 2020). Moreover, the number of identified studies for dementia and Parkinson’s disease was too small to allow for reliable pooling of the data. Based on the limited evidence available the observed benefits on cognitive, physical and psychosocial function for these populations appear to be more modest, however, the heterogeneous intervention and control conditions included across a small set of studies limits firm conclusions.

The results from the network meta-analysis showed that the cognitive effects of simultaneous and sequential training exceeded all control conditions apart from cognitive training alone, and that simultaneous training was likely to be the most beneficial approach. Systematic reviews indicate that both physical (Firth et al., 2018) and cognitive training (van Balkom et al., 2020) lead to positive neurobiological changes with potential therapeutic relevance. The therapeutic premise of combined interventions is that physical and cognitive activities might influence brain plasticity through distinct and complementary pathways, whereby physical exercise induces physiological changes (e.g., upregulation of brain-derived neurotrophic factor, stimulation of hippocampal neurogenesis) which, in turn, facilitates the experience-dependent neuroplastic effects of cognitive engagement (Fissler et al., 2013; Kempermann et al., 2010). Due to the transient release of neurotrophic factors following physical exercise (Knaepen et al., 2010), it has been suggested that the physical and cognitive activity should be conducted in close temporal proximity to achieve maximal benefit (Fissler et al., 2013), with potential order effect (Nilsson et al., 2020). This provides a potential mechanistic account for the observed cognitive benefits of simultaneous training, but the evidence base is still inconclusive and likely dependent on exercise intensity. Therefore, the optimal combination method remains an open question in the field, warranting head-to-head trials aiming to optimise efficacy and adherence while taking into account the feasibility of practical implementation.

Moreover, our results confirm that both simultaneous and sequential training augment the cognitive effects of physical exercise alone. While this is consistent with findings from previous meta-analyses (Gheysen et al., 2018; Zhu et al., 2016), the current network meta-analysis focusing specifically on RCTs and with moderate confidence ratings for the relevant comparisons provides more robust evidence on the benefits of enriching physical exercise with structured cognitive challenge in order to maximize cognitive gains.

For physical function, the results from the network meta-analysis showed comparable efficacy of simultaneous training, sequential training and physical exercise alone. This suggests that combining physical exercise with cognitive training, even in a simultaneous design, does not come at the cost of reduced efficacy of the physical intervention component. In contrast, the cognitive and physical benefits of exergaming appear to be smaller than those of simultaneous and sequential training. One potential explanation for this observation is that the cognitive and physical demands of exergaming might be lower compared to more traditional cognitive and physical training. Our results contrast the medium-sized cognitive effect found for active videogames by Stanmore et al. (2017) and are closer to those reported in a re-analysis by Sala et al. (2019). While gamification of health interventions, such as exergaming, may facilitate intervention design characteristics that motivate behaviour change (Johnson et al., 2016), ensuring that the cognitive and physical demands of exergaming interventions match those of more traditional physical exercise and cognitive training interventions seems imperative.

From a clinical and practical perspective, the cognitive and physical improvements observed following simultaneous training are encouraging, since this is a time- and resource-effective approach for targeting multiple risk factors for cognitive and functional decline. Sequential physical and cognitive training interventions generally require high-frequency training, which may induce stress and fatigue reactions that counteract training effects (Lampit et al., 2014; Zhu et al., 2016). Additionally, it has been suggested that simultaneous training may have positive effects on training enjoyment, as compared to sequential training (McEwen et al., 2018) and physical exercise (Eggenberger et al., 2015a), although this issue needs further clarification. Based on the results from the sensitivity analyses, we found no evidence to suggest that simultaneous training is less feasible or efficacious for cognition in MCI compared to cognitively healthy older adults, indicating that this approach can be successfully implemented also for those with mildly impaired cognition. An important avenue for future research is to delineate the specific intervention characteristics of simultaneous training, such as physical and cognitive training content, dose and frequency that are the most important for training effectiveness, to inform practical implementation.

### 4.2 Strengths and limitations

To the best of our knowledge, this is the first systematic review and multivariate meta-analysis of RCTs investigating combined cognitive physical and cognitive training across a wide range of outcomes of relevance for health and functional independence in older age, thus providing a comprehensive synthesis of the current evidence-base for combined interventions. Moreover, our use of a network meta-analytical approach facilitates simultaneous treatment comparisons and improves precision of the effect size estimates (Salanti, 2012) and the current review is substantially larger than the previous network meta-analysis in the field (Bruderer-Hofstetter et al., 2018). This allows firmer conclusions regarding the comparative effectiveness and relative rankings of the different types of combined interventions, and evaluation of the confidence in the findings. Nevertheless, some limitations should be addressed. First, although we used stringent inclusion criteria to ensure that the interventions were reasonably similar across studies, the included treatments nevertheless consisted of a variety of physical exercise (e.g., aerobic, strength, mobility and balance) and cognitive training (e.g., computerized or non-computerized, targeting single or multiple cognitive domains, videogames) elements. This suggests that further dismantling of the specific cognitive and physical components that make up an effective combined intervention should be explored; this could be achieved through component network meta-analysis (Rücker et al., 2020b). Nevertheless, there was no substantial statistical heterogeneity or inconsistency in the results of the network meta-analyses and the certainty of the evidence was moderate to high for most comparisons of cognitive outcomes. In contrast, the confidence in the results of the network meta-analysis for physical outcomes was low to very low for several of the comparisons, mainly owing to concerns regarding imprecision in the effect estimates and within-study bias. Thus, while these initial results are encouraging, the low certainty of the evidence for physical outcomes suggests that the relative treatment estimates may change as a consequence of future, high-quality research. Second, we did not identify enough studies for dementia and Parkinson’s disease to allow for reliable pooling of the data. Consequently, further research is needed to establish the potential benefits and comparative efficacy of combined interventions for individuals with more prominent cognitive and/or physical impairment. Third, investigation of potential effect modifiers such as exercise type, intensity or training frequency was not feasible under the network meta-analysis framework due to insufficient number of studies within each node. Although a larger meta-analysis of physical exercise did not find evidence to support exercise modalities as a mediator of cognitive outcomes (Falck et al., 2019), such effects are likely and would be informative for future research and practice in the field. Additionally, the relevance of intervention settings and provision of social support should be explored; these considerations might be particularly relevant for individuals with cognitive and functional impairment. Fourth, trials that recruited community dwelling elderly participants did not always provide information on global cognitive status (e.g., Mini-Mental State Examination scores), and the methods for defining MCI varied across studies. Thus, some heterogeneity within these groups is likely. Future trials should make efforts to describe the cognitive status of participants and use established diagnostic criteria for MCI. Finally, this meta-analysis was restricted to investigation of post-intervention effects and the effects of combined interventions in the medium-long term should be explored.

### 4.3 Conclusions

Combined cognitive and physical training, delivered either simultaneously or sequentially, is efficacious in promoting both cognitive and physical health in older age, at least in the short term. These benefits were observed in cognitively healthy older adults, as well as for people with MCI. Simultaneous training showed comparable cognitive effects as sequential training and cognitive training alone, while also achieving similar physical improvements as physical exercise, suggesting this is a resource-effective approach for targeting multiple dementia risk factors. Exergaming interventions might incorporate elements from simultaneous training in game design to augment their cognitive and physical efficacy. More research is needed to establish whether cognitive and physical improvements following combined interventions translate into improved everyday function and well-being, particularly so for individuals with cognitive and functional impairment, as well as what specific components make up an effective combined intervention and what regimens are needed in order to maintain long-term gains.

## Data Availability

All underlying data are appended to the manuscript.

## Acknowledgements

We thank Sahar Aghajari for help with data entry and the authors of primary studies for providing data and advice. MLM was funded by a scholarship from Dementia Australia. ATW is funded by a grant from the Australian National Health and Medical Research Council (NHMRC GNT1171313). AL is funded by a CR Roper Fellowship from the University of Melbourne.

## Appendix

## Appendix A. Search strategy (Ovid)

1. (cognitive training or brain training or attention training or reasoning training or memory training or mental training or mental skills training or neurocognitive training).mp
2. (cognitive exercise or brain exercise or memory exercise or attention exercise or reasoning exercise).mp
3. (cognitive stimulation or memory stimulation or memory enhanc$ or cognitive enhanc$).mp
4. (cognitive rehabilitat$ or cognitive remediation or cognitive restructur$).mp
5. (cognitive activit$ or mental activit$).mp
6. (speed and processing and training).mp
7. (mnemonic$ or method of loci).mp
8. (video game$ or videogame$ or wii or computer game$ or virtual reality).mp
9. (cognitive intervention$ or neurocognitive intervention$ or neuropsychological intervention$).mp
10. exercis$.mp
11. sport$.mp
12. exp Exercise/
13. exp Physical fitness/
14. (aerobic exercis$ or aerobic train$ or aerobic fitness or aerobic program$).mp
15. (resistance exercis$ or resistance train$ or anaerobic exercis$ or anaerobic train$ or resistance program$).mp
16. (physical or aerobic or endurance or cardiorespiratory or cardiovascular or resistance or strength).mp
17. (bicycl$ or bike rid$ or bicycle rid$).mp
18. (multimodal or multidomain or multicomponent or multi-modal or multi-domain or multi-component or dual task or dual-task or tai chi or taiji or tai chi chuan or danc$).mp
19. (exergam$ or active video game$ or active videogame$ or kinect or active play or interactive video).mp
20. (cognitive adj2 physical).mp
21. (cognition or cognitive or memory or executive function$ or executive control or attention or visuospatial or processing speed or language).mp
22. (older adults or elder$ or senior$ or adult$ or older or ag?ing).mp
23. (mild cognitive impairment or mildly cognitively impaired or Parkins$ or dement$ or alzheimer$).mp
24. 1 or 2 or 3 or 4 or 5 or 6 or 7 or 8 or 9
25. 10 or 11 or 12 or 13 or 14 or 15 or 16 or 17
26. 22 or 23
27. 24 and 25
28. 18 or 19 or 20
29. (27 or 28) and 21 and 26

## Appendix B. Classification of cognitive and physical outcomes

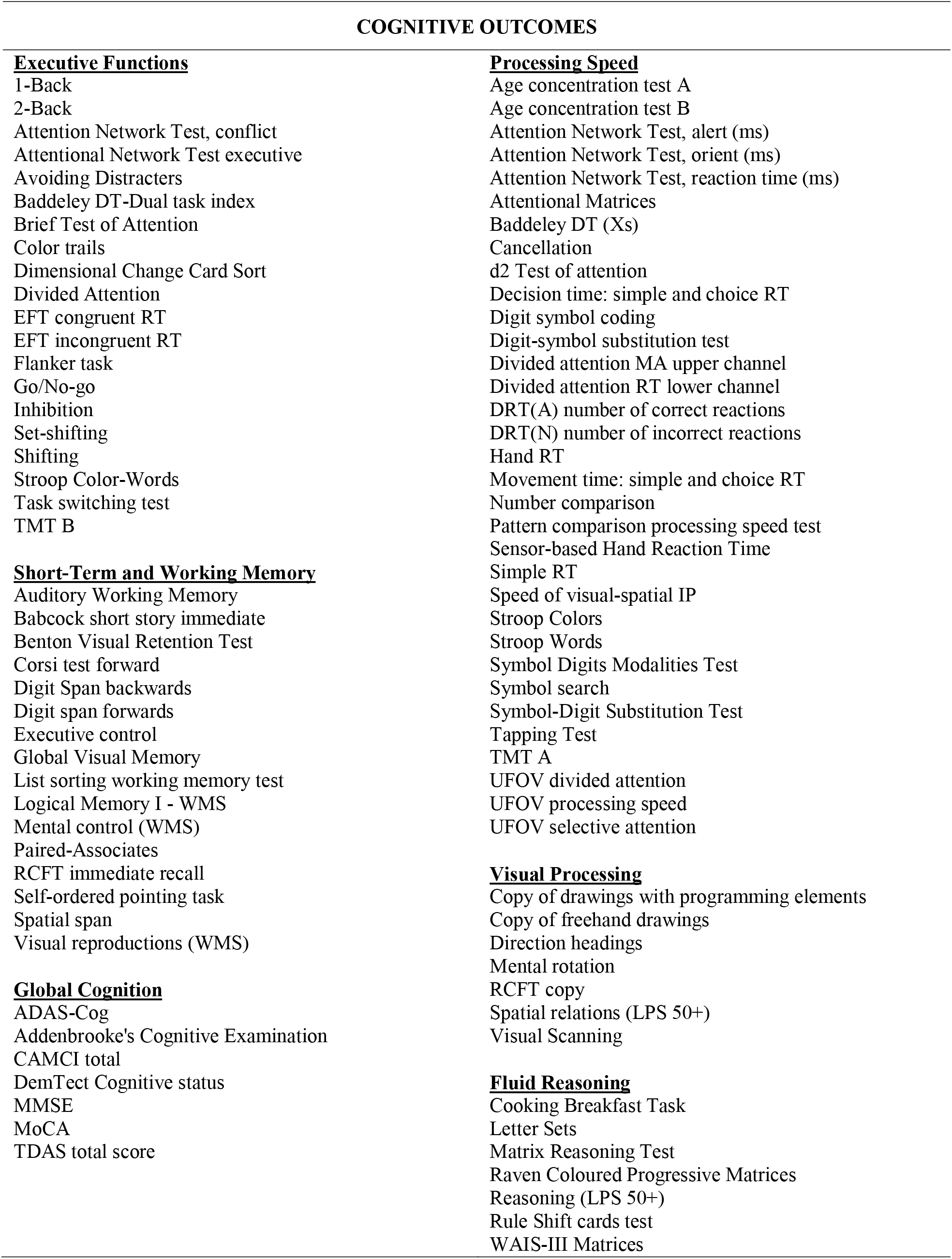
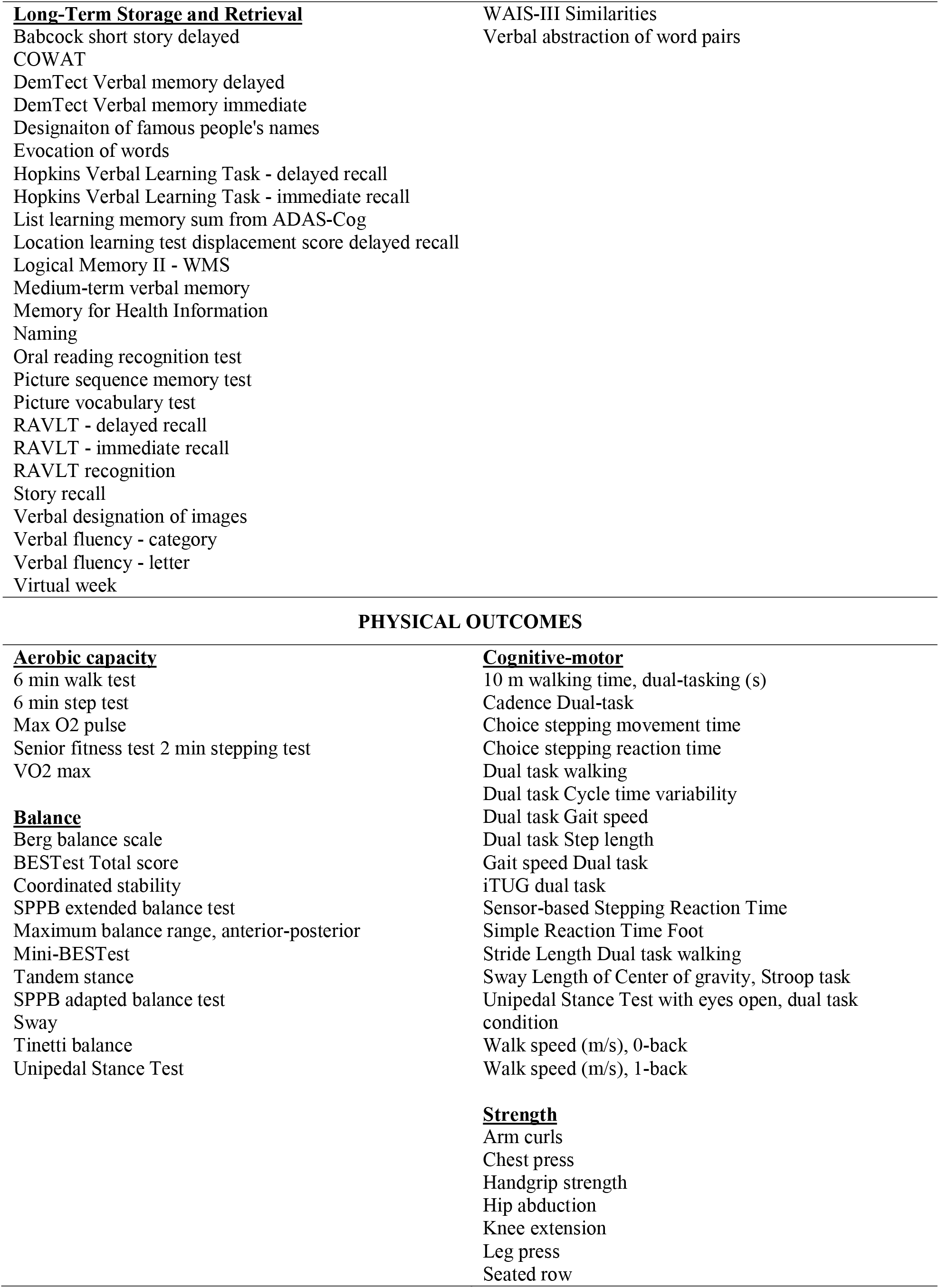
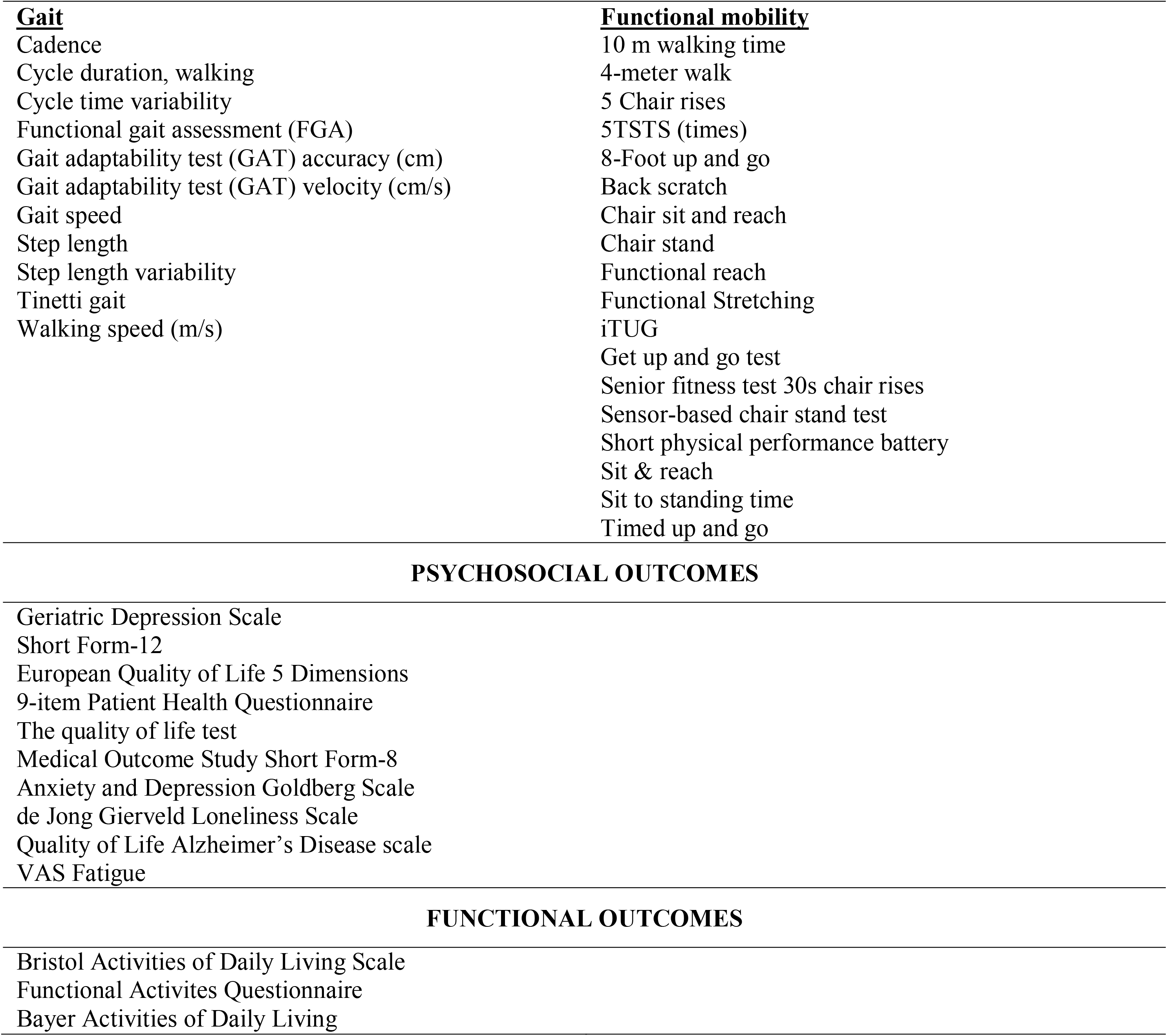

## Appendix C. List of excluded studies. (Note: a study could be ineligible for multiple reasons but appear only once in this table.)

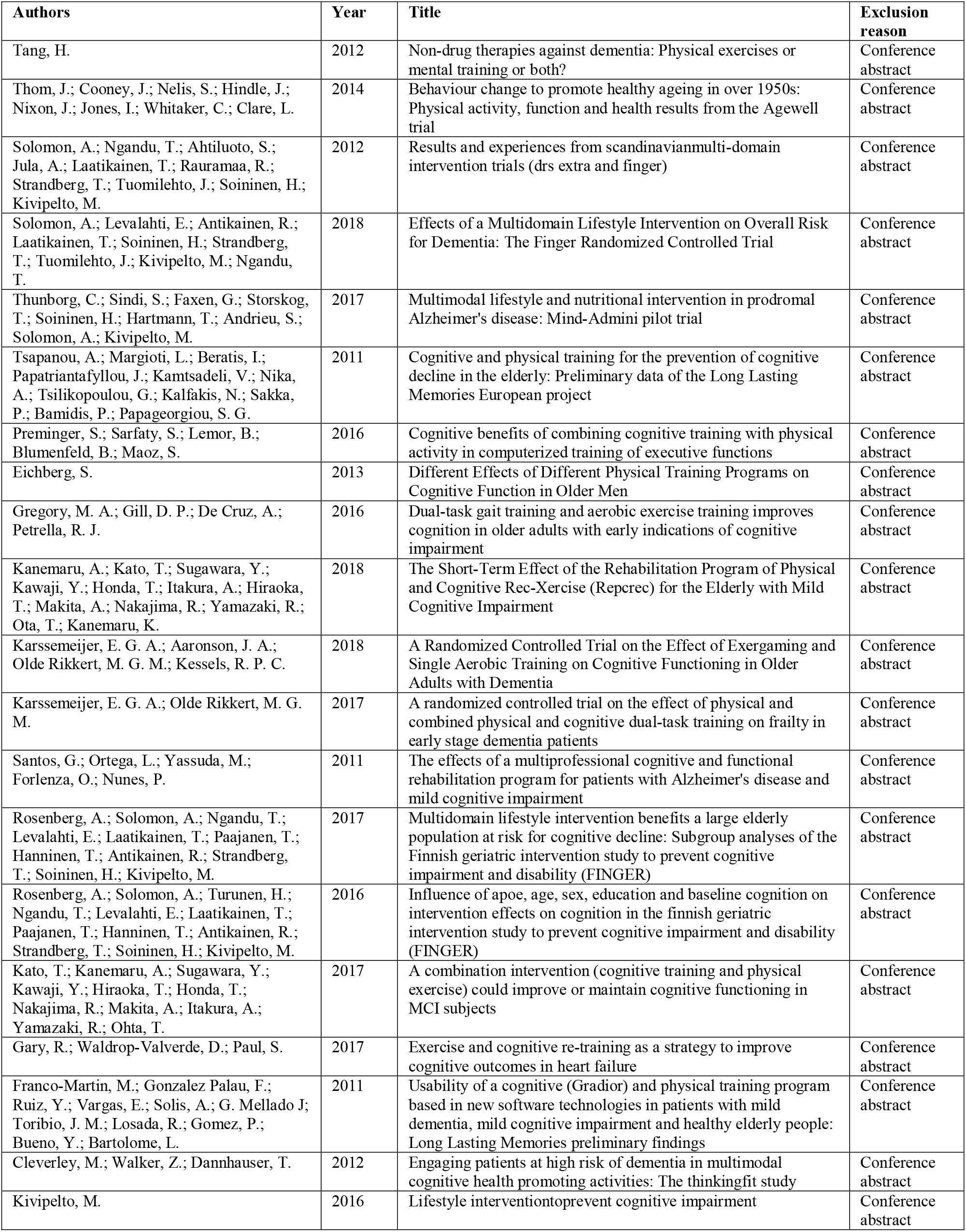
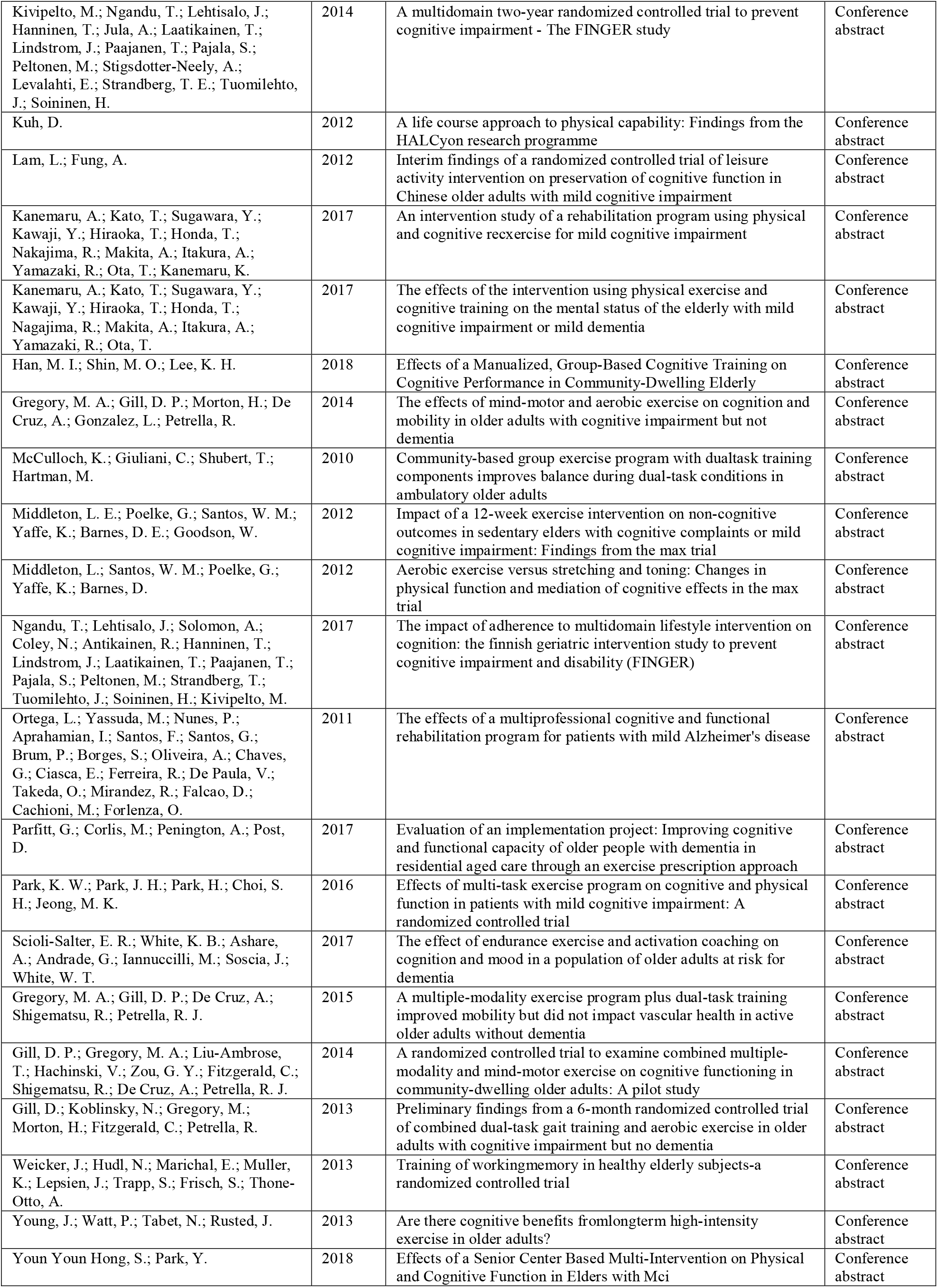
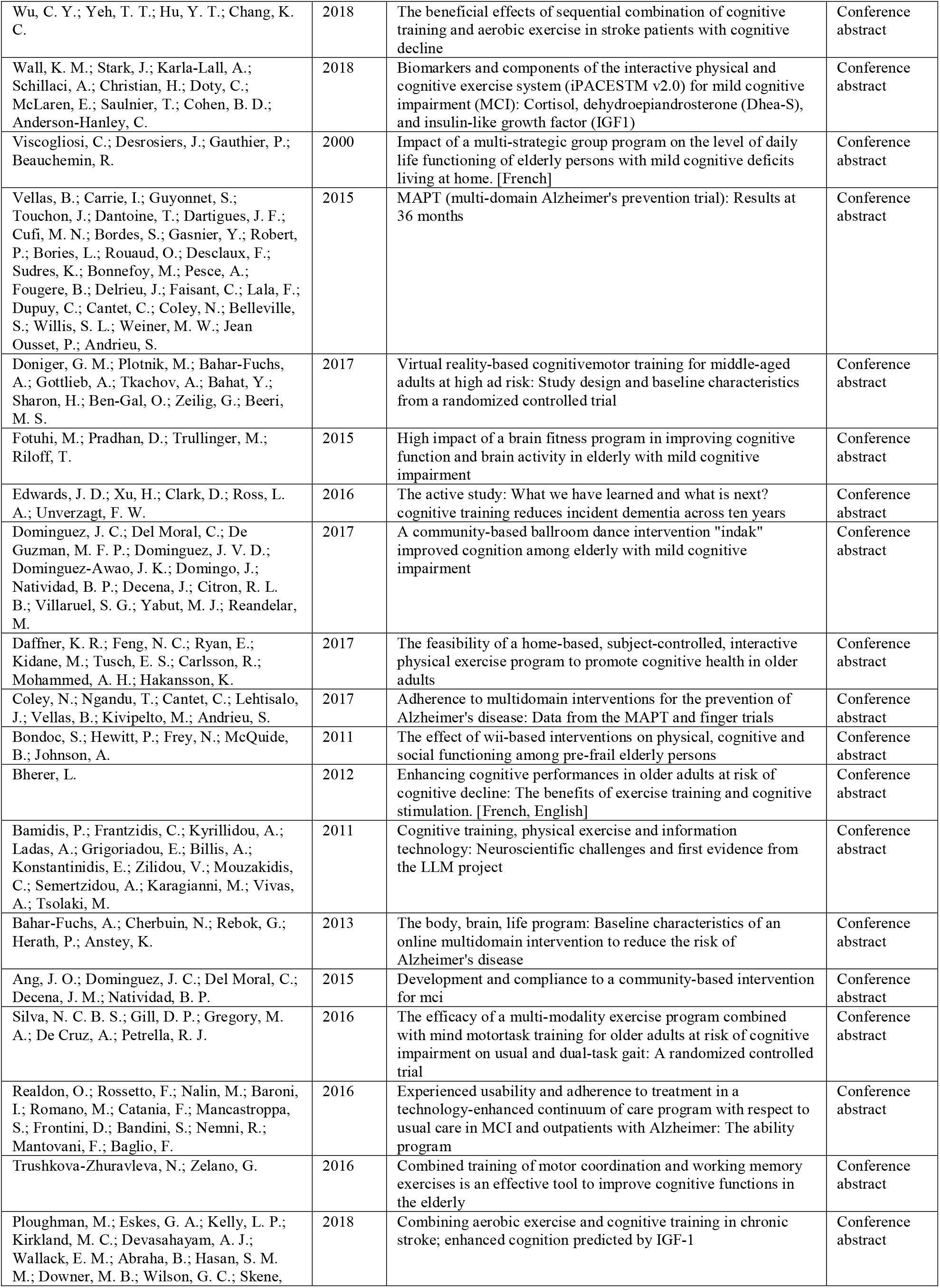
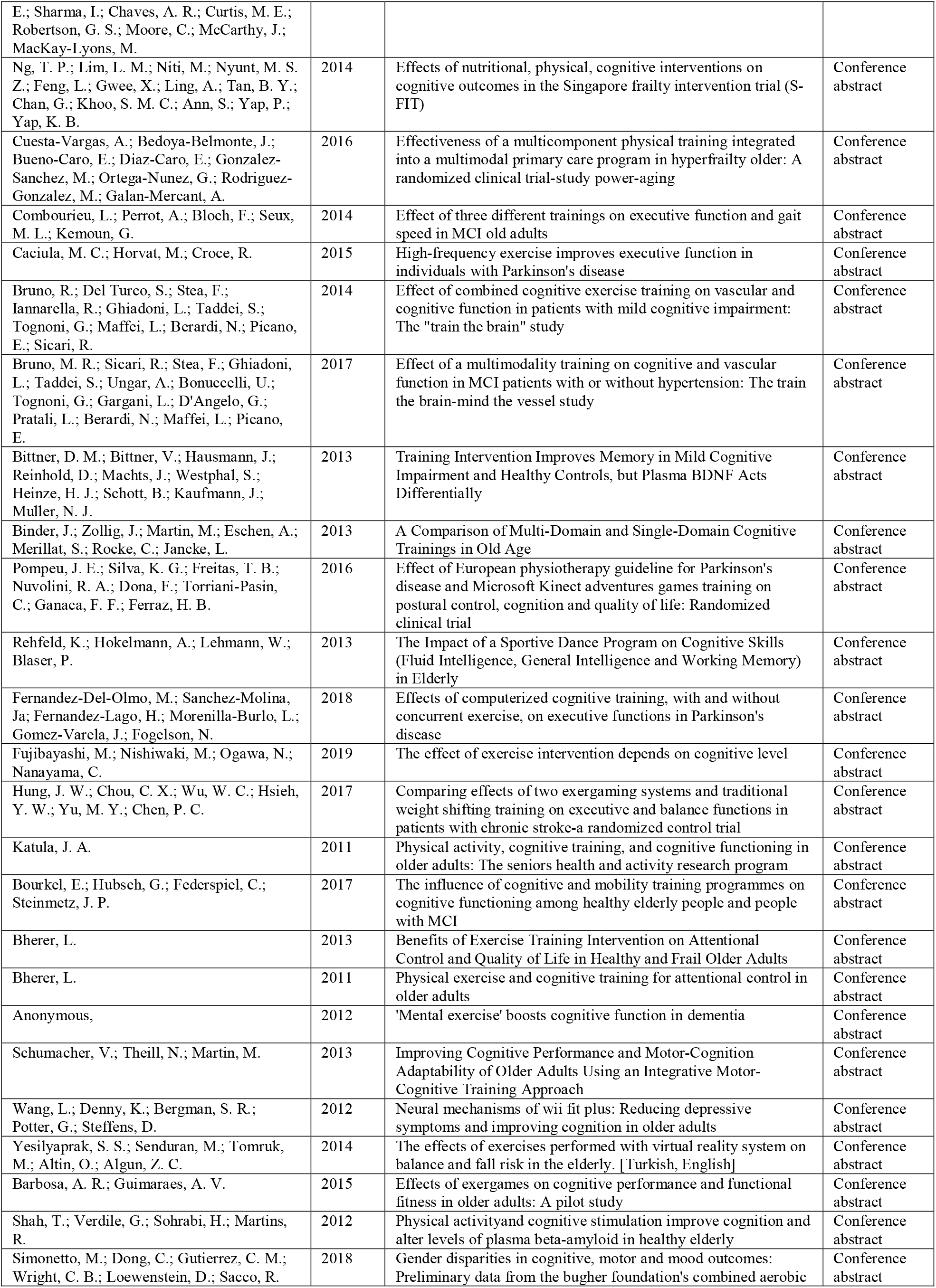
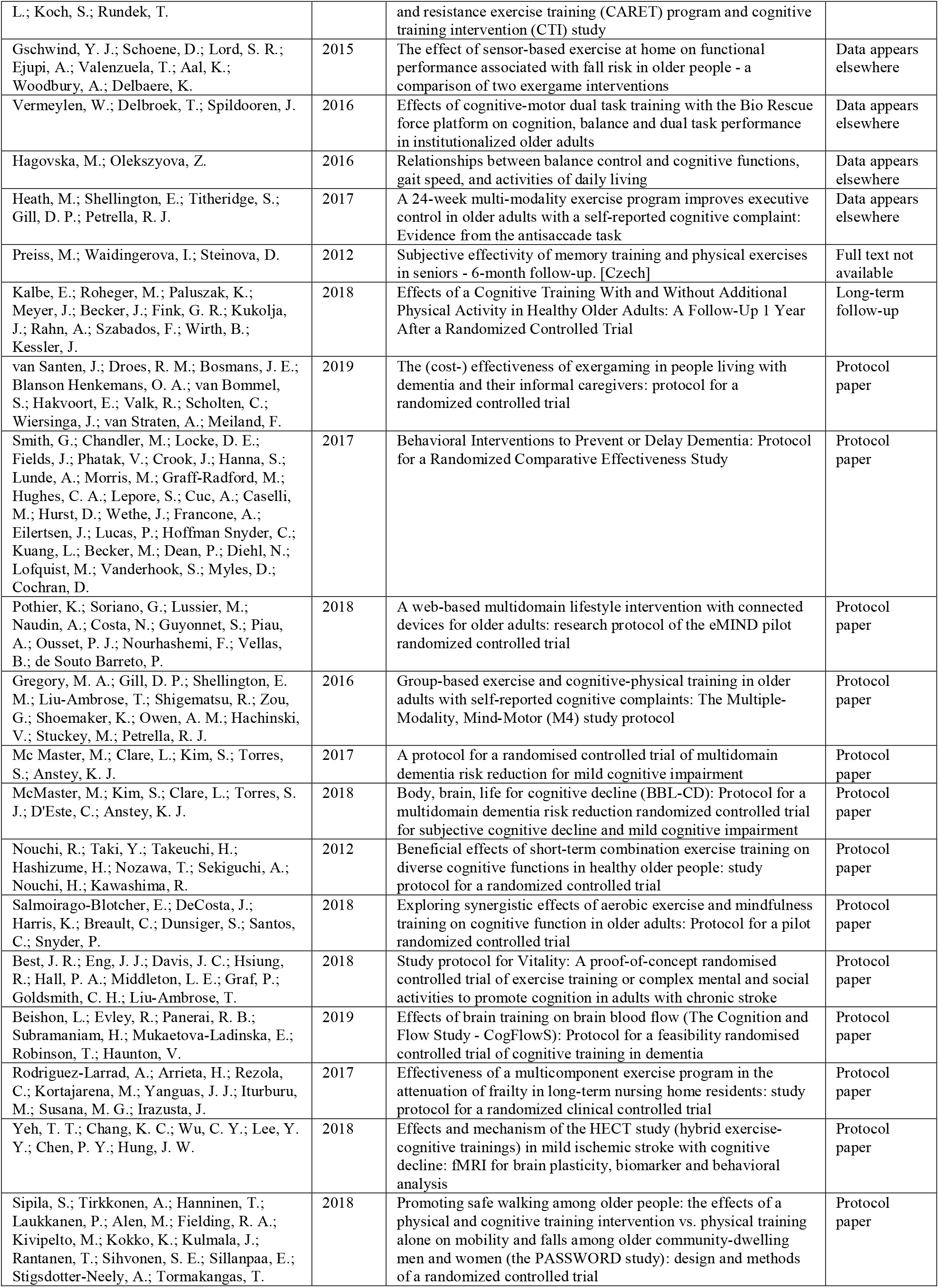
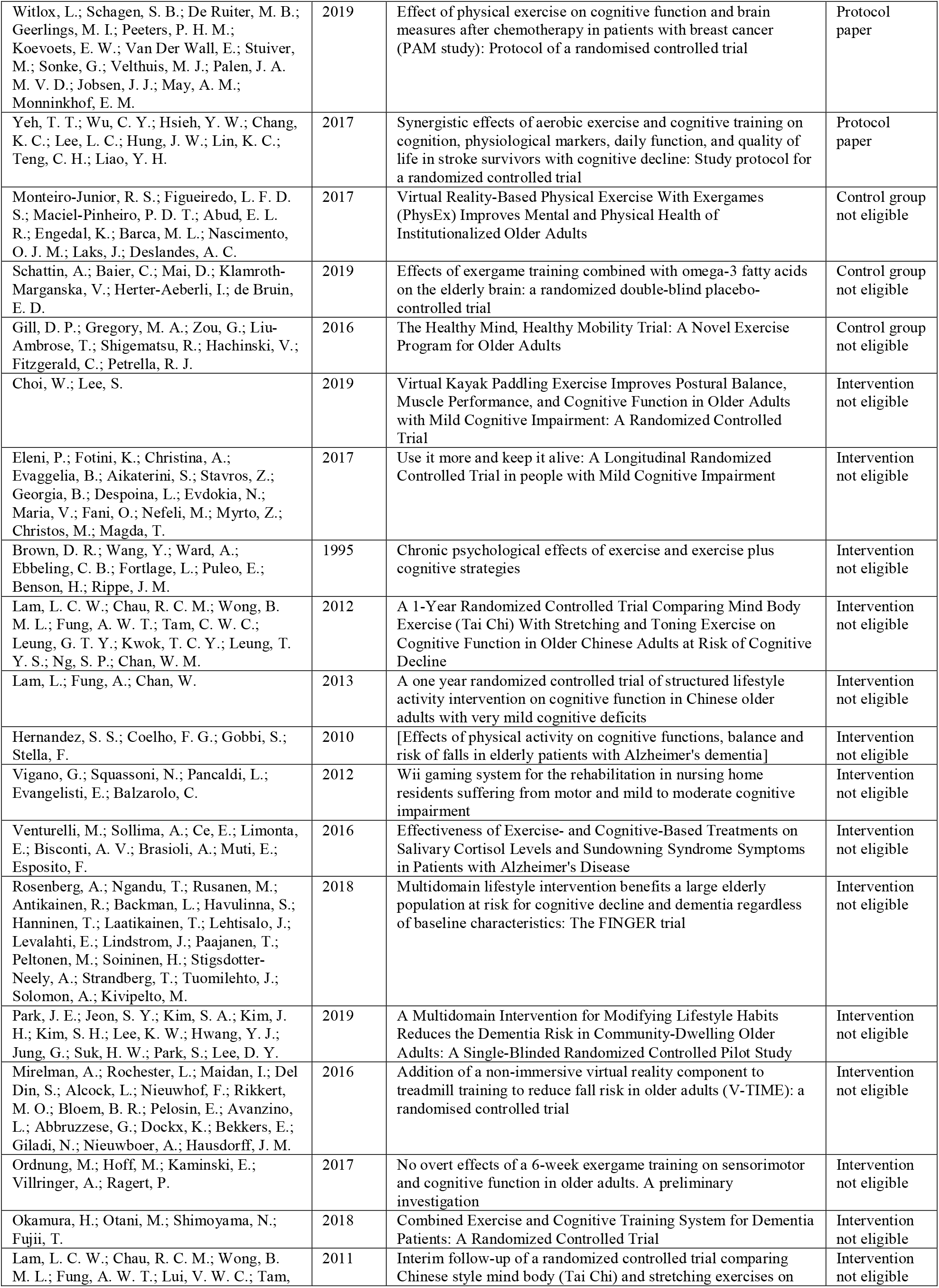
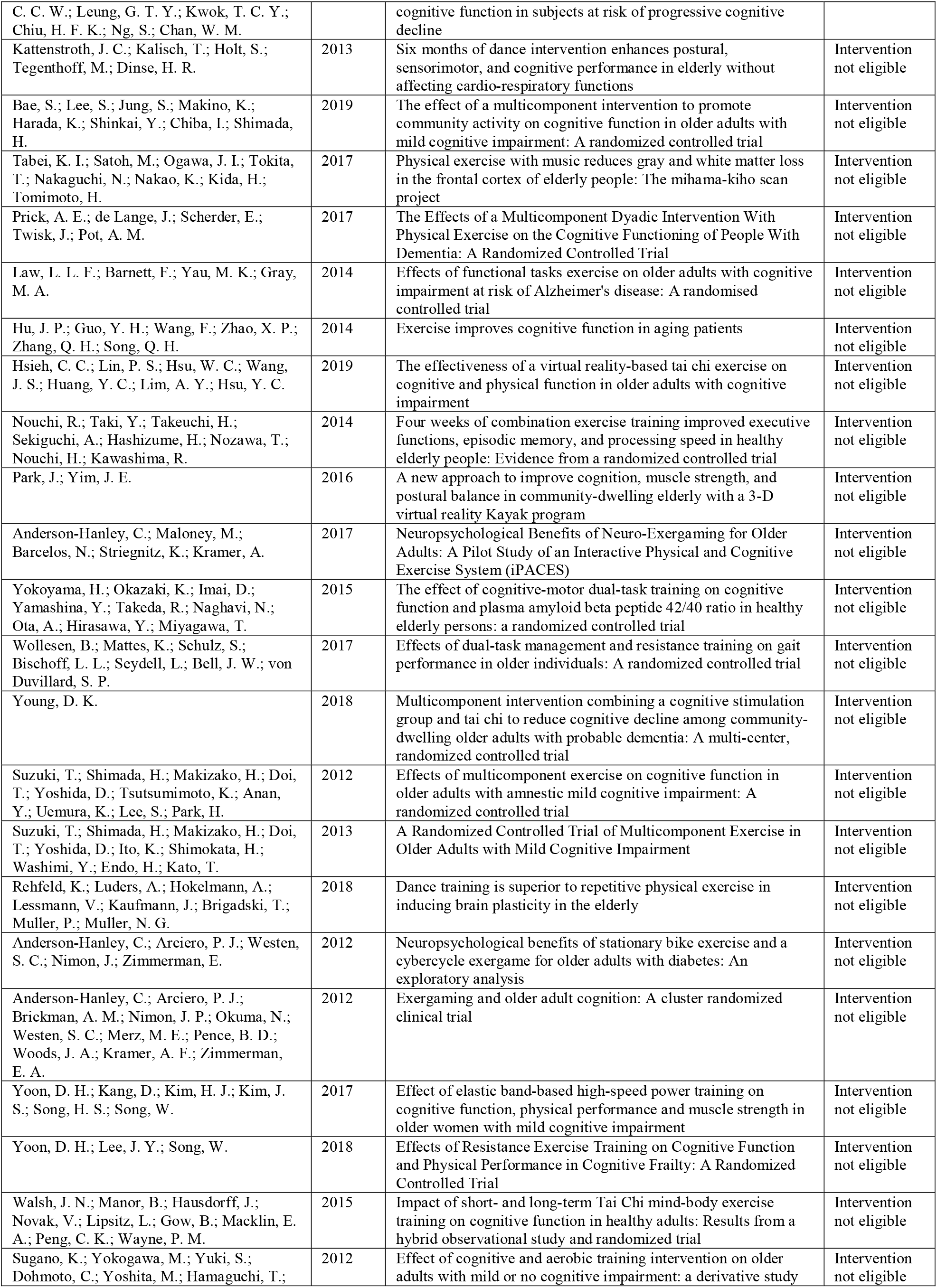
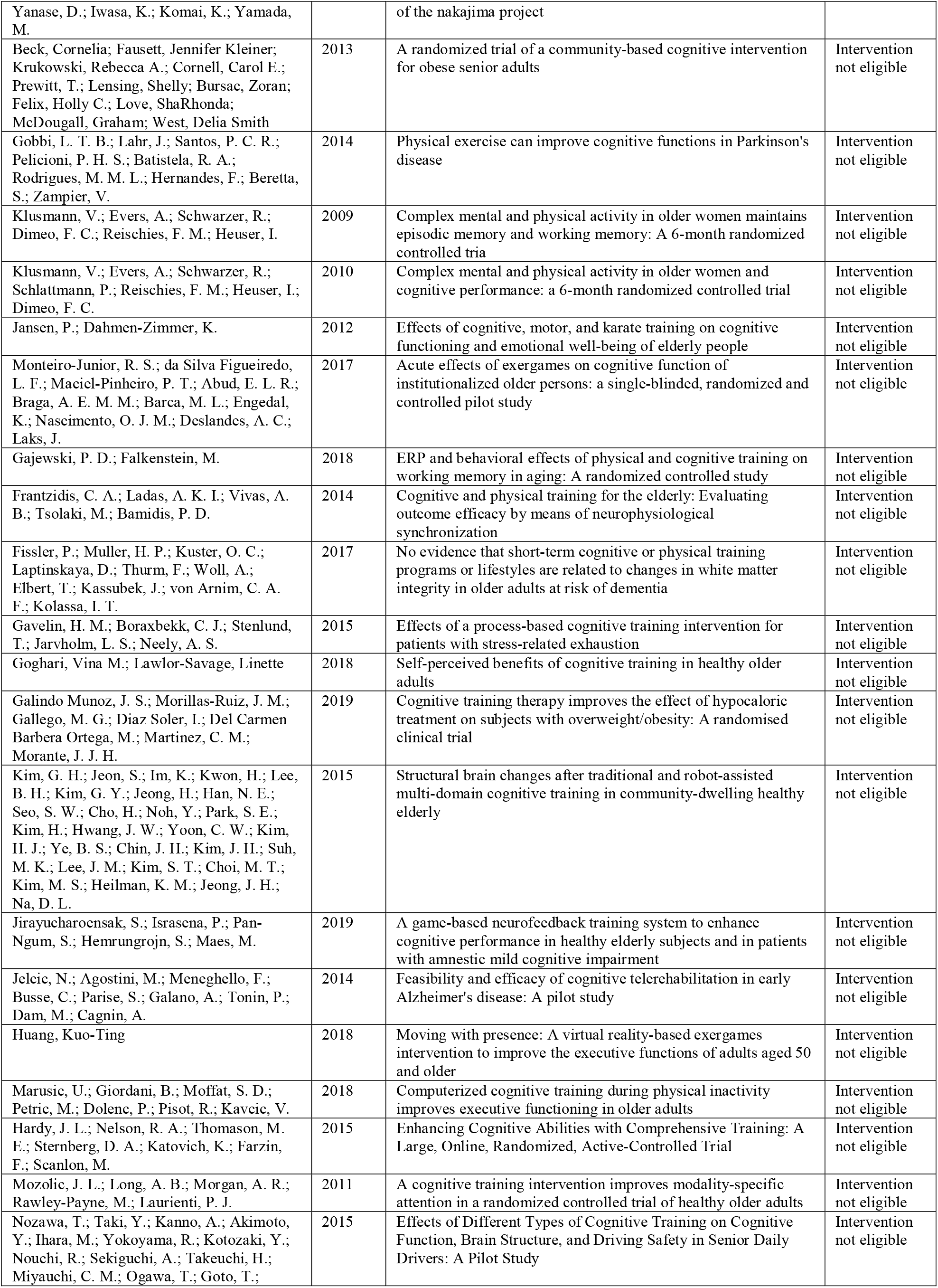
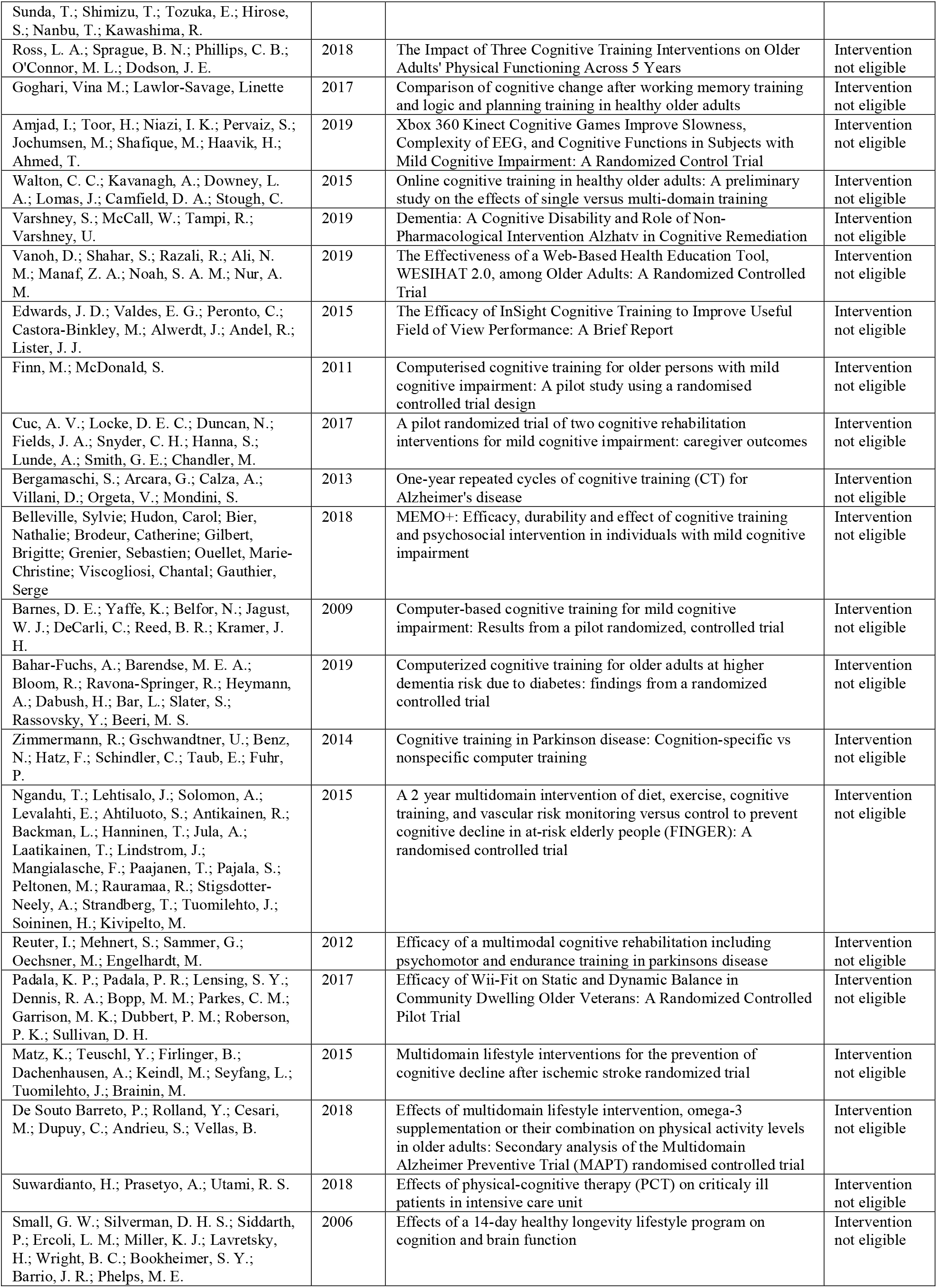
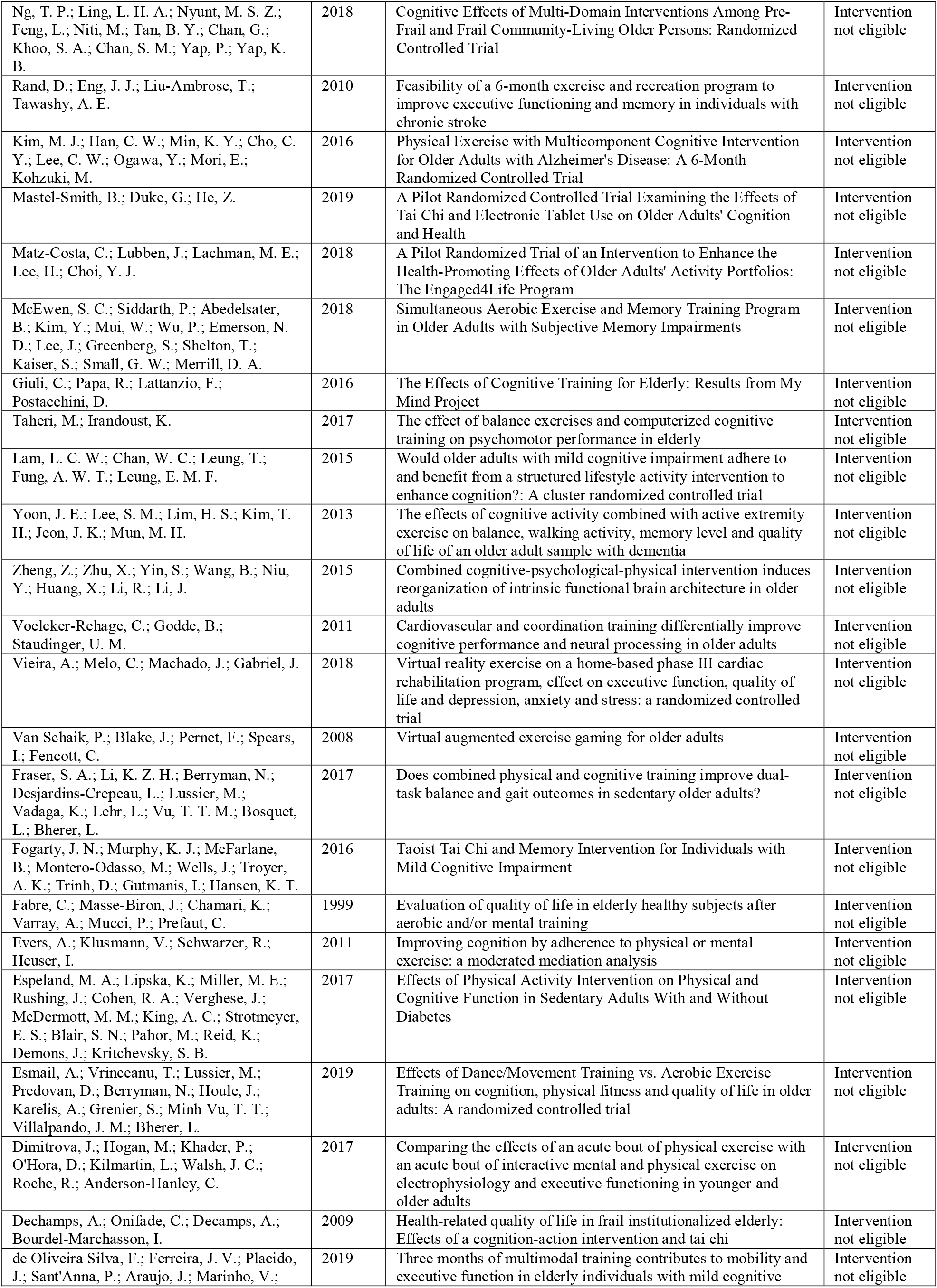
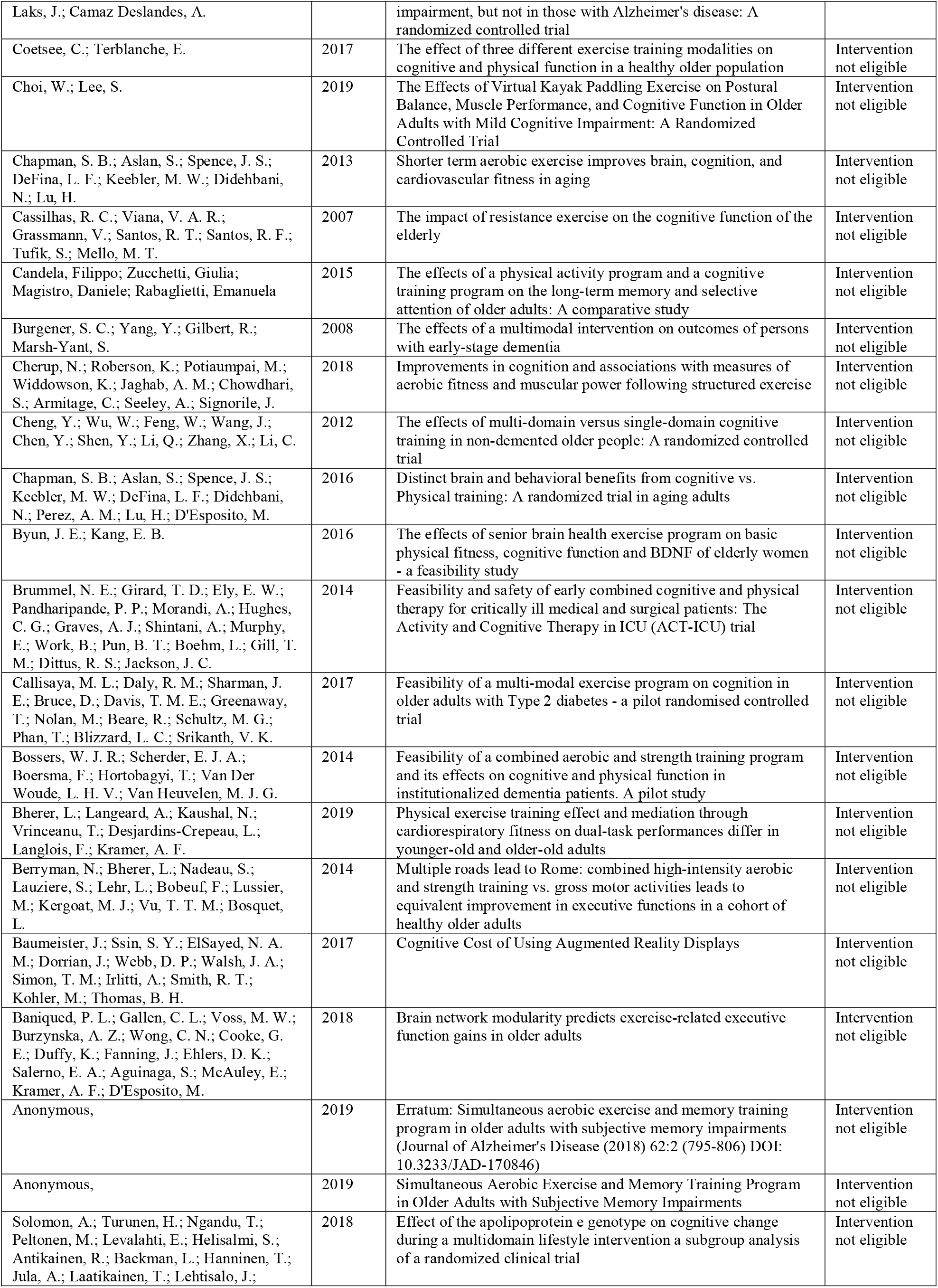
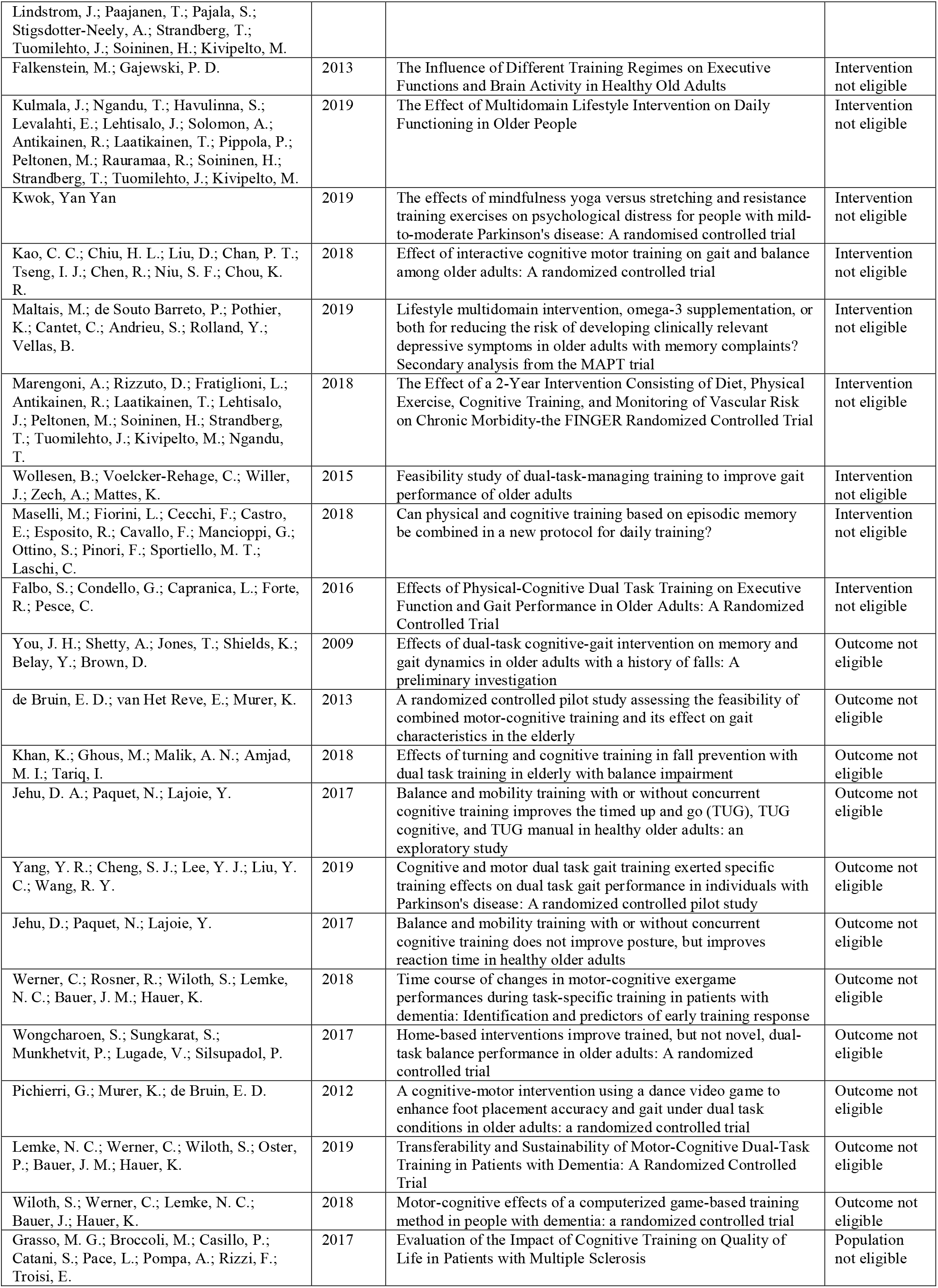
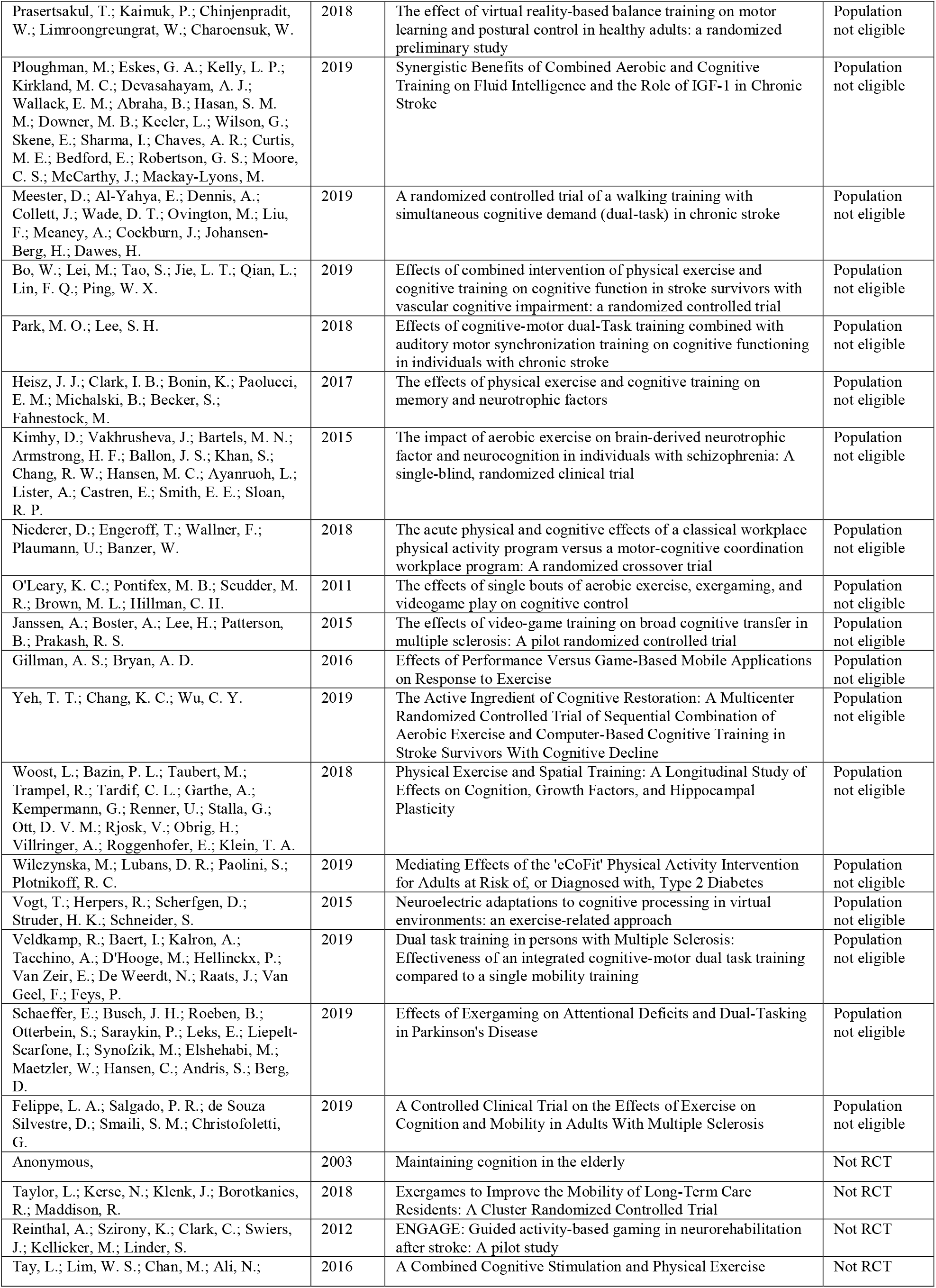
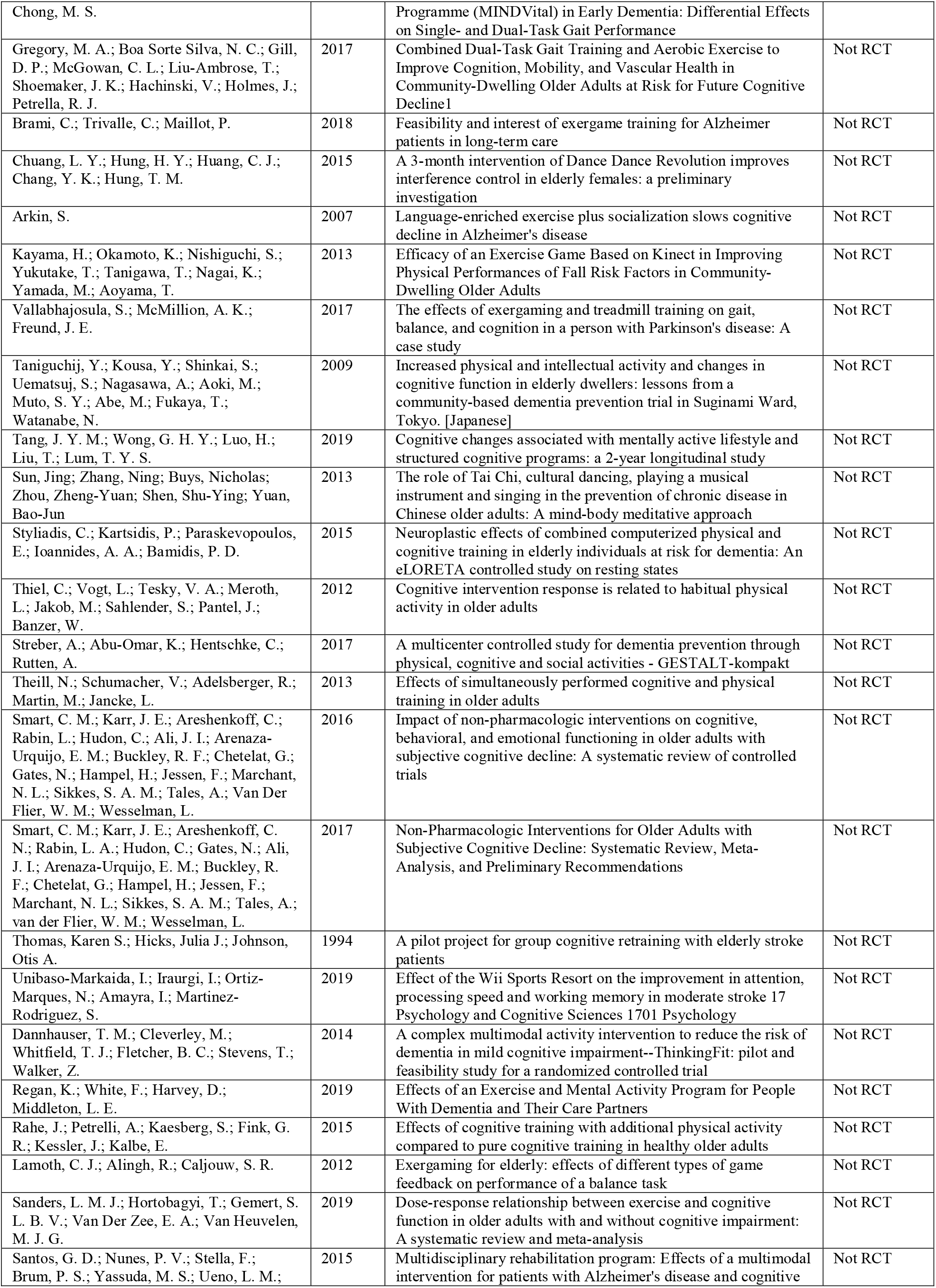
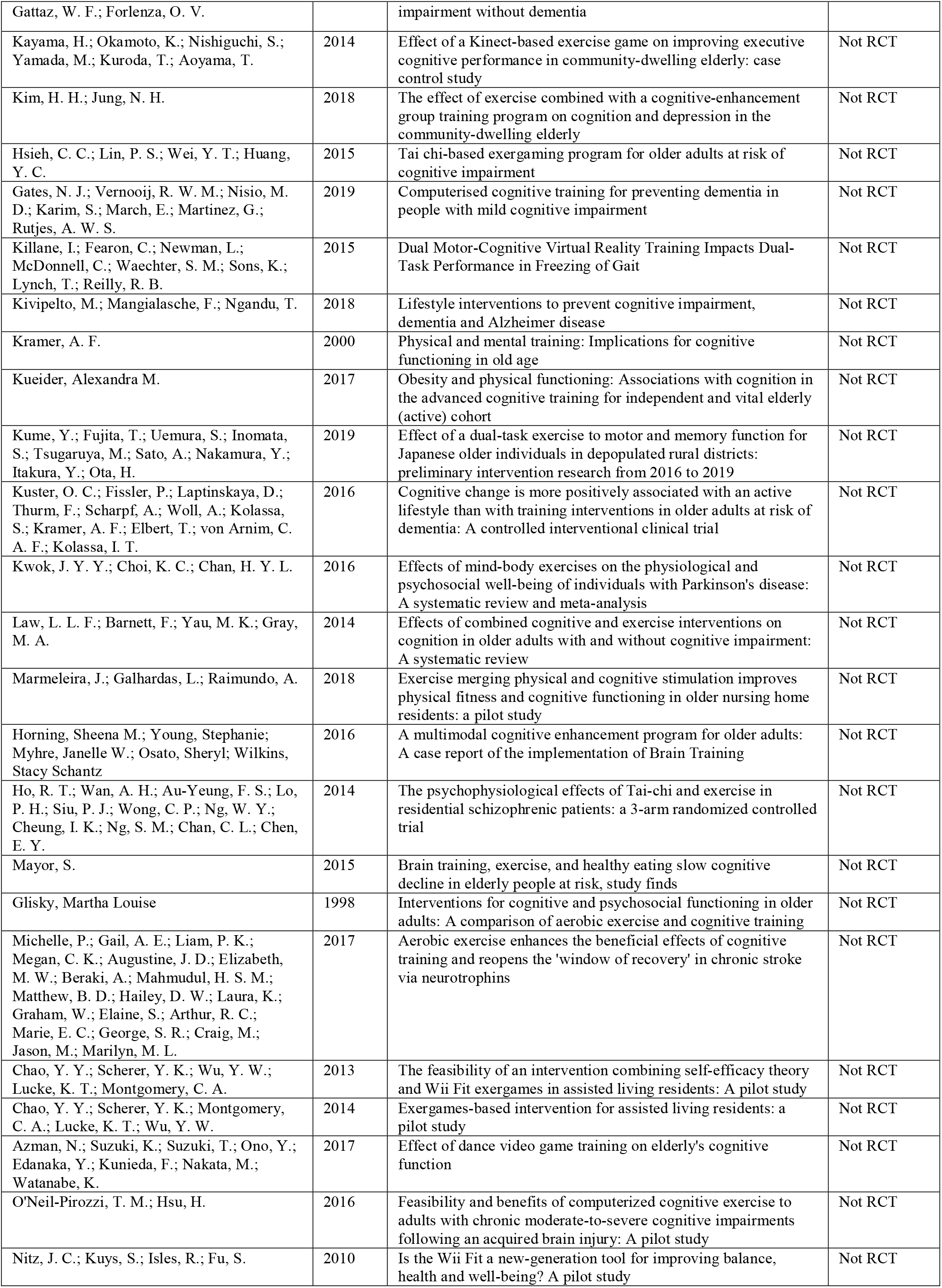
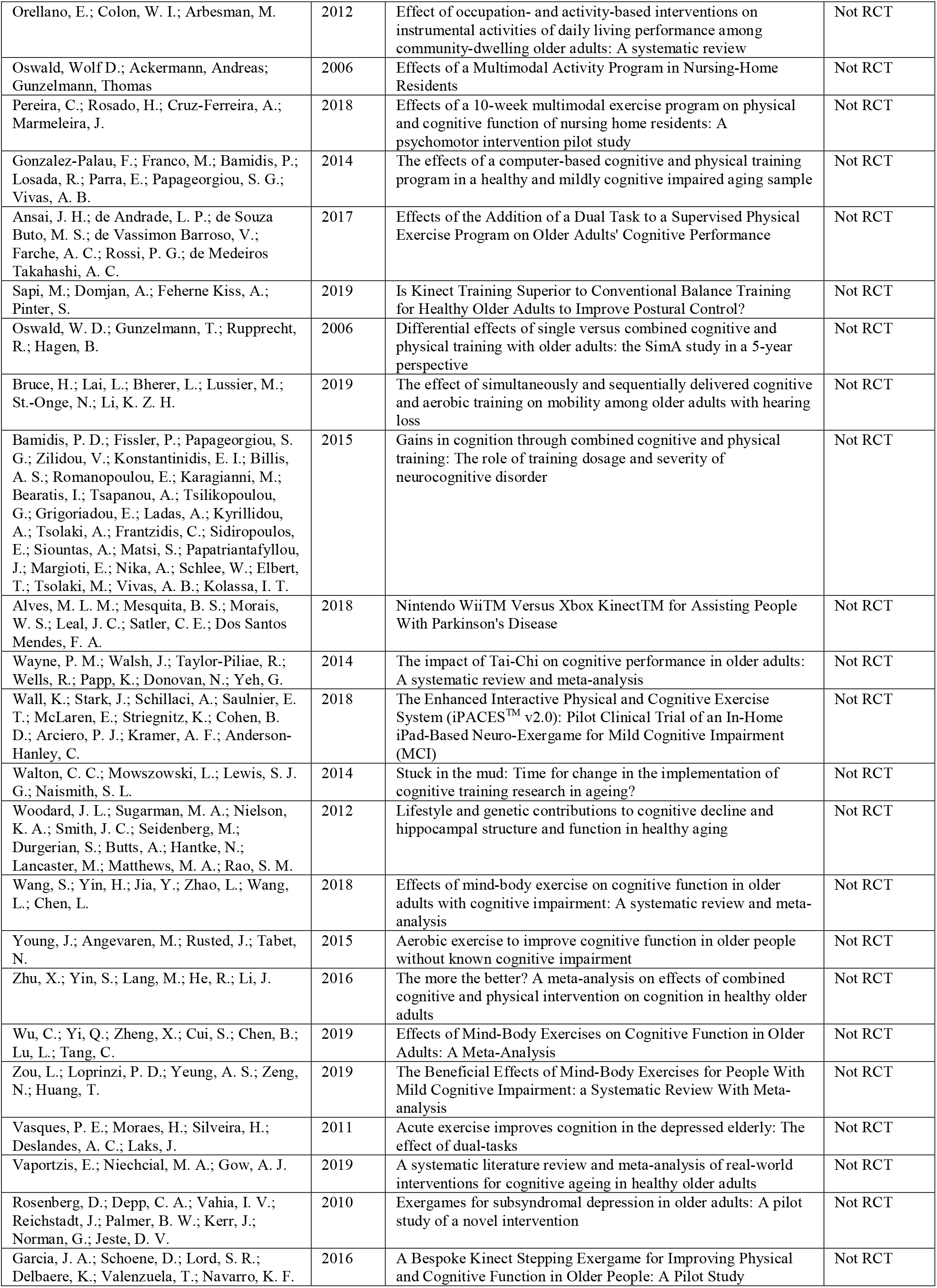
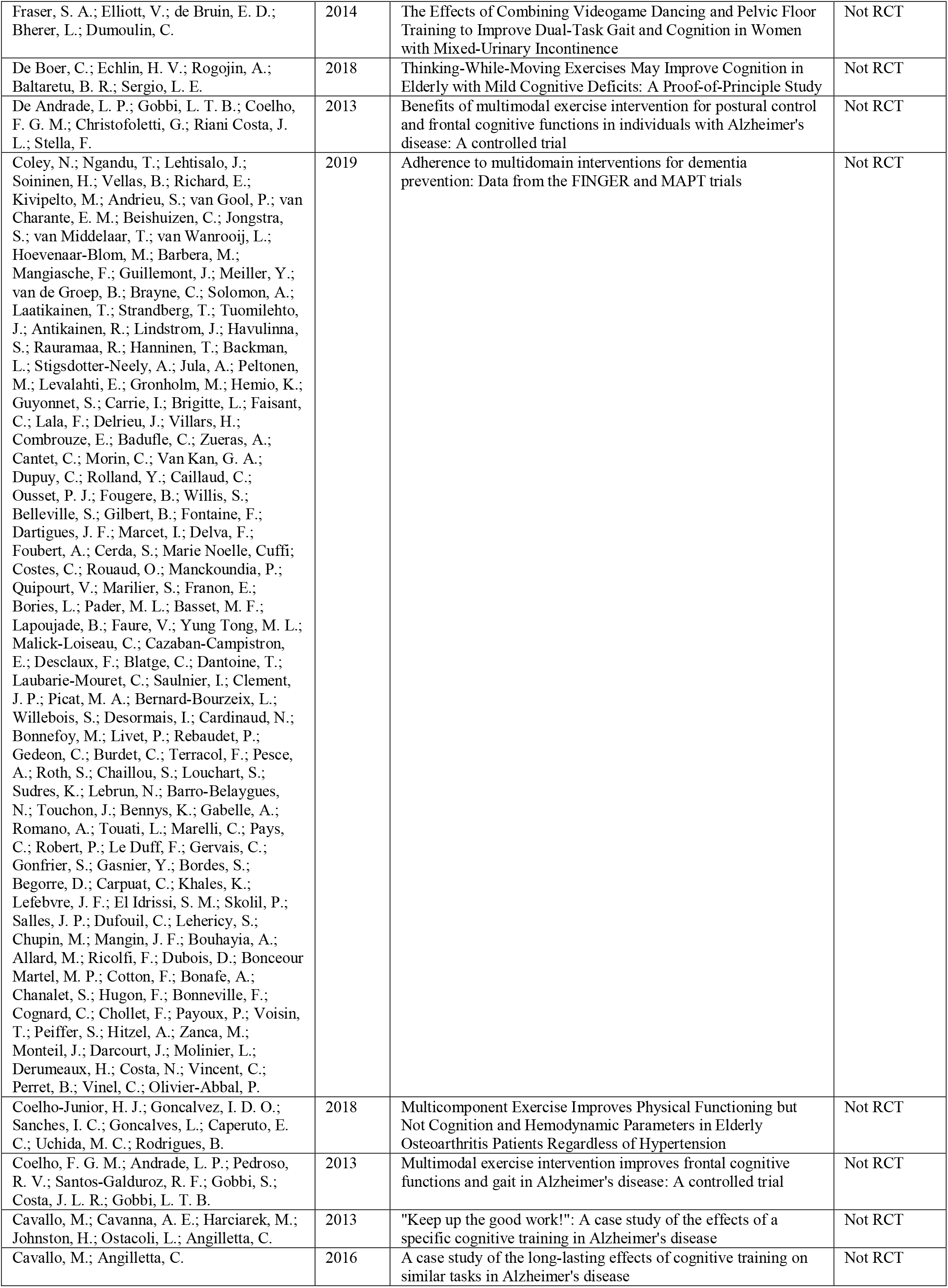
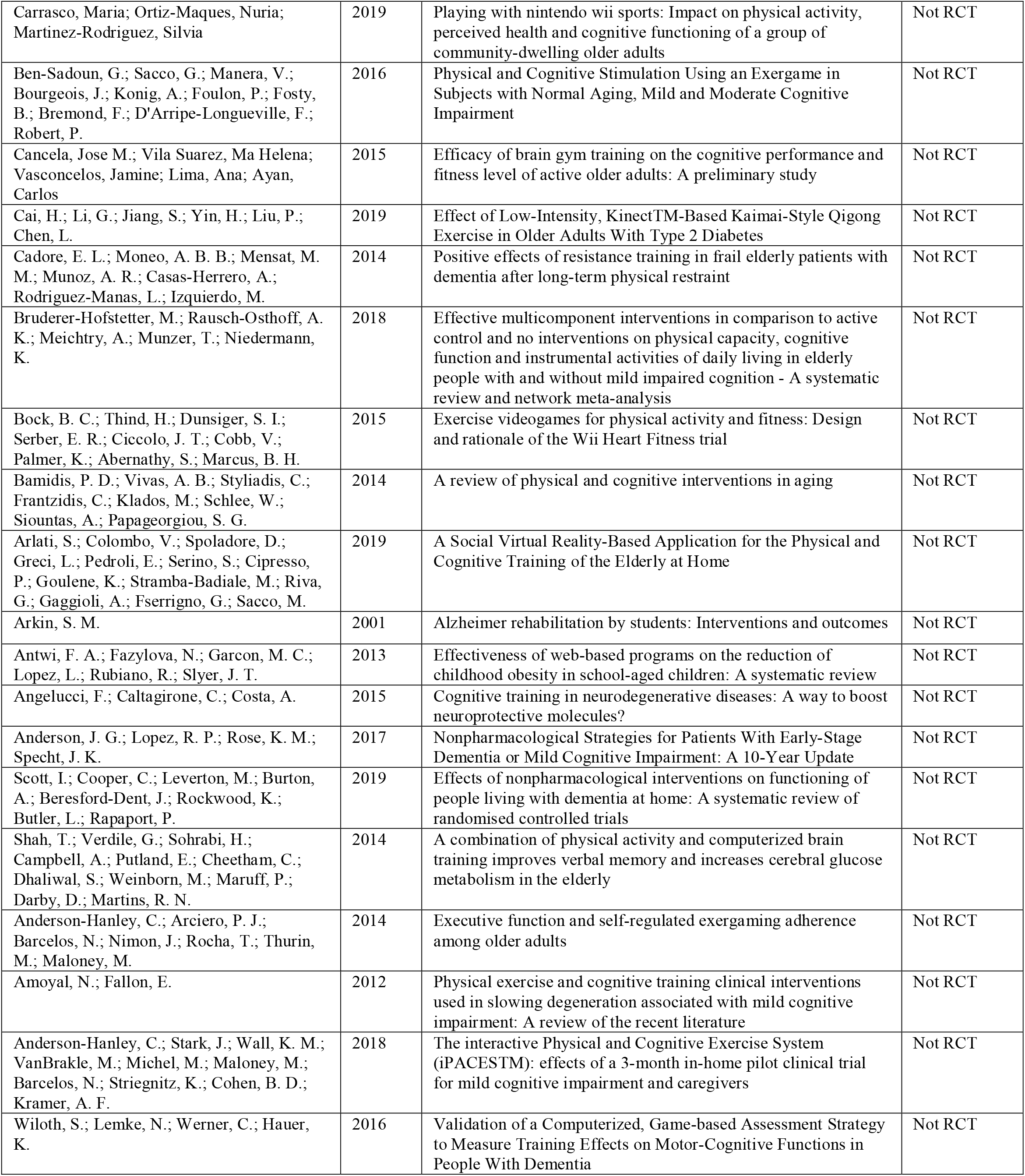

## Appendix D. Risk of bias within individual studies

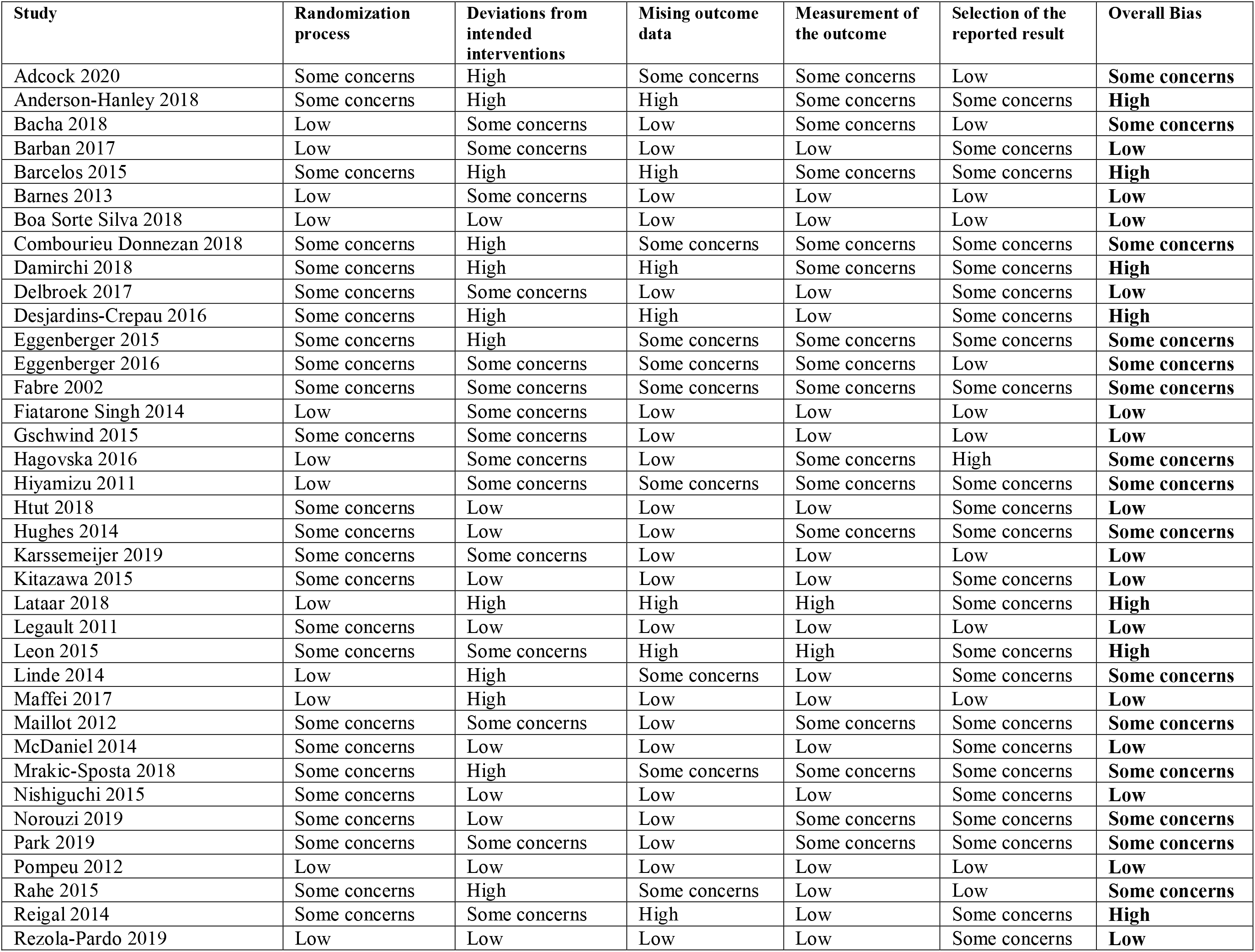
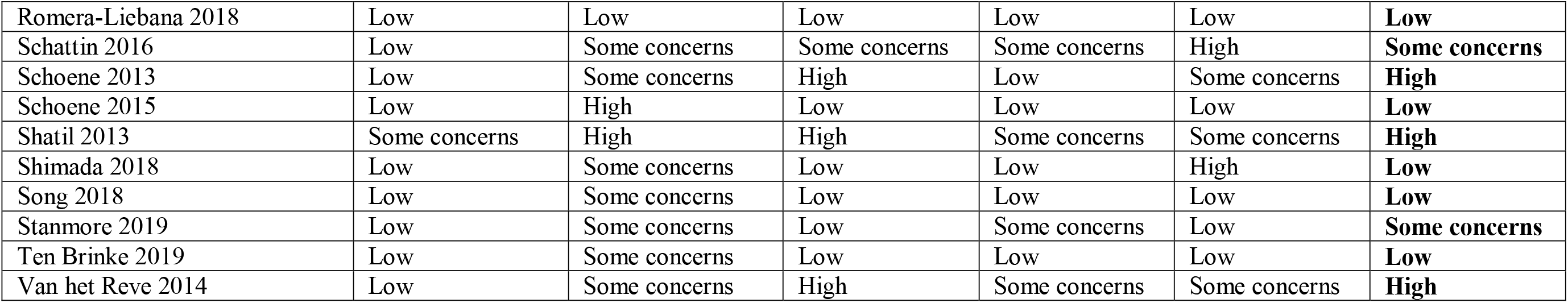

## Appendix E. Funnel plots of cognitive, physical and psychosocial outcomes

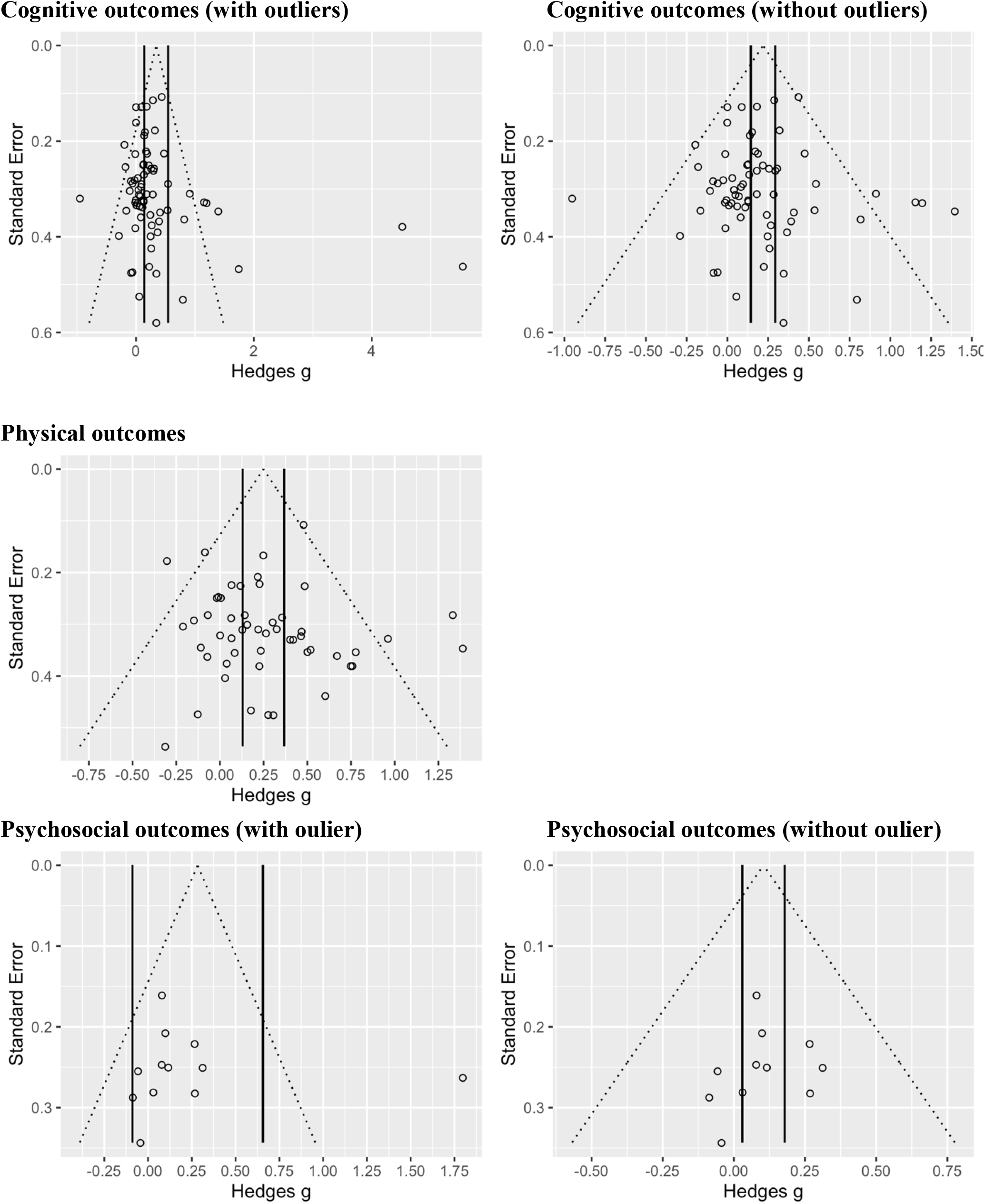

## Appendix F. Results of individual studies and comparisons: Overall cognition

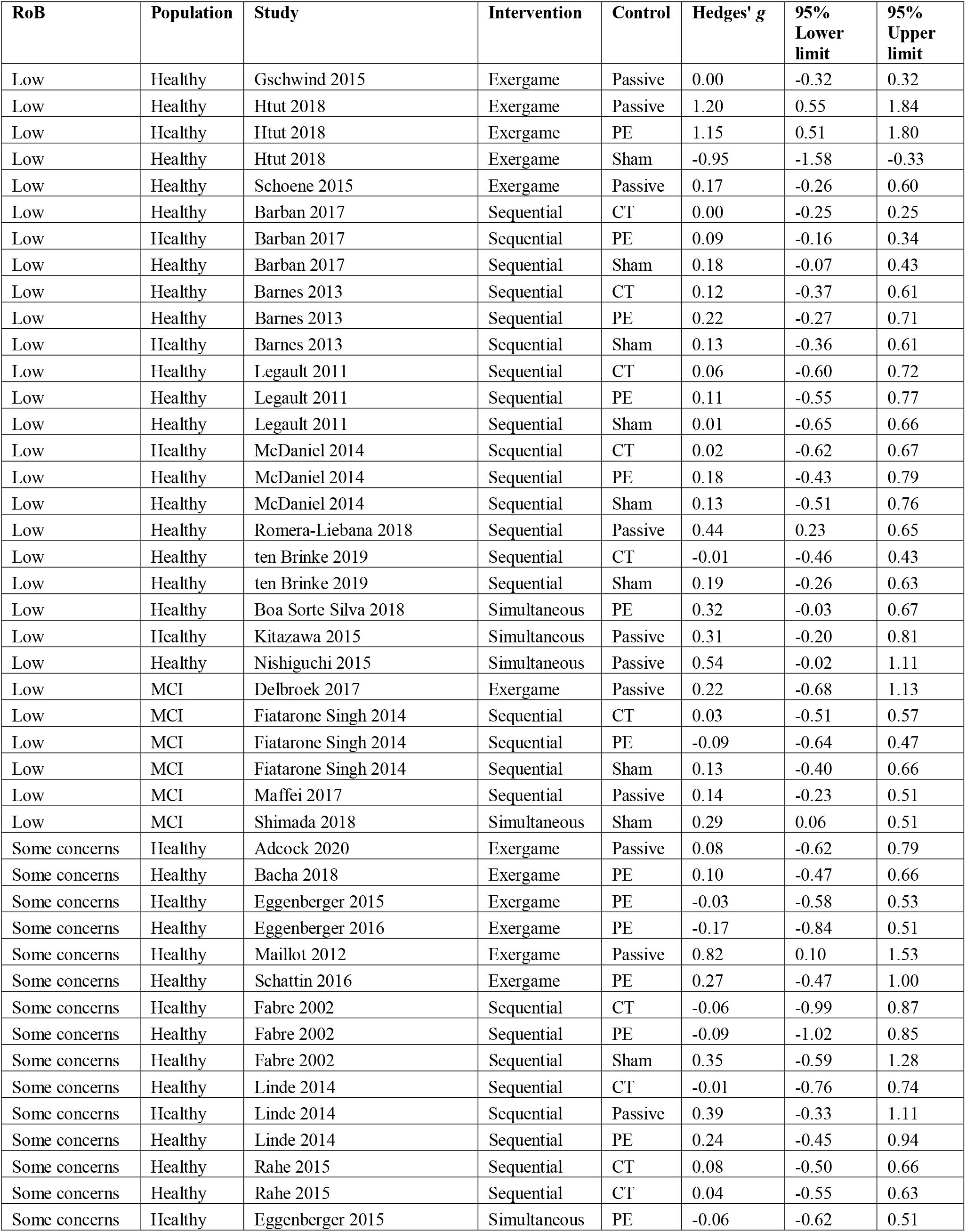
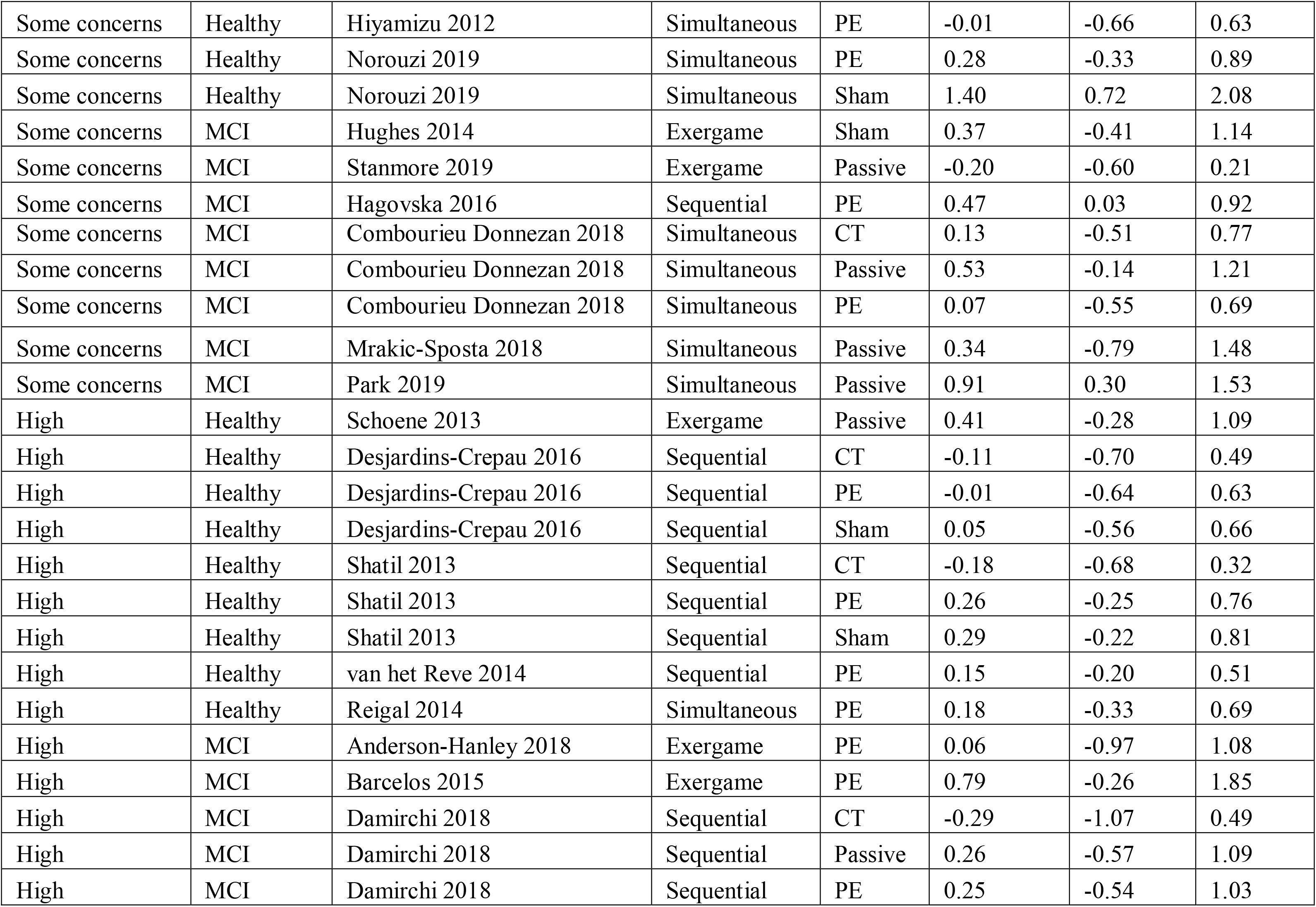

## Appendix G. Results of individual studies and comparisons: Overall physical outcomes

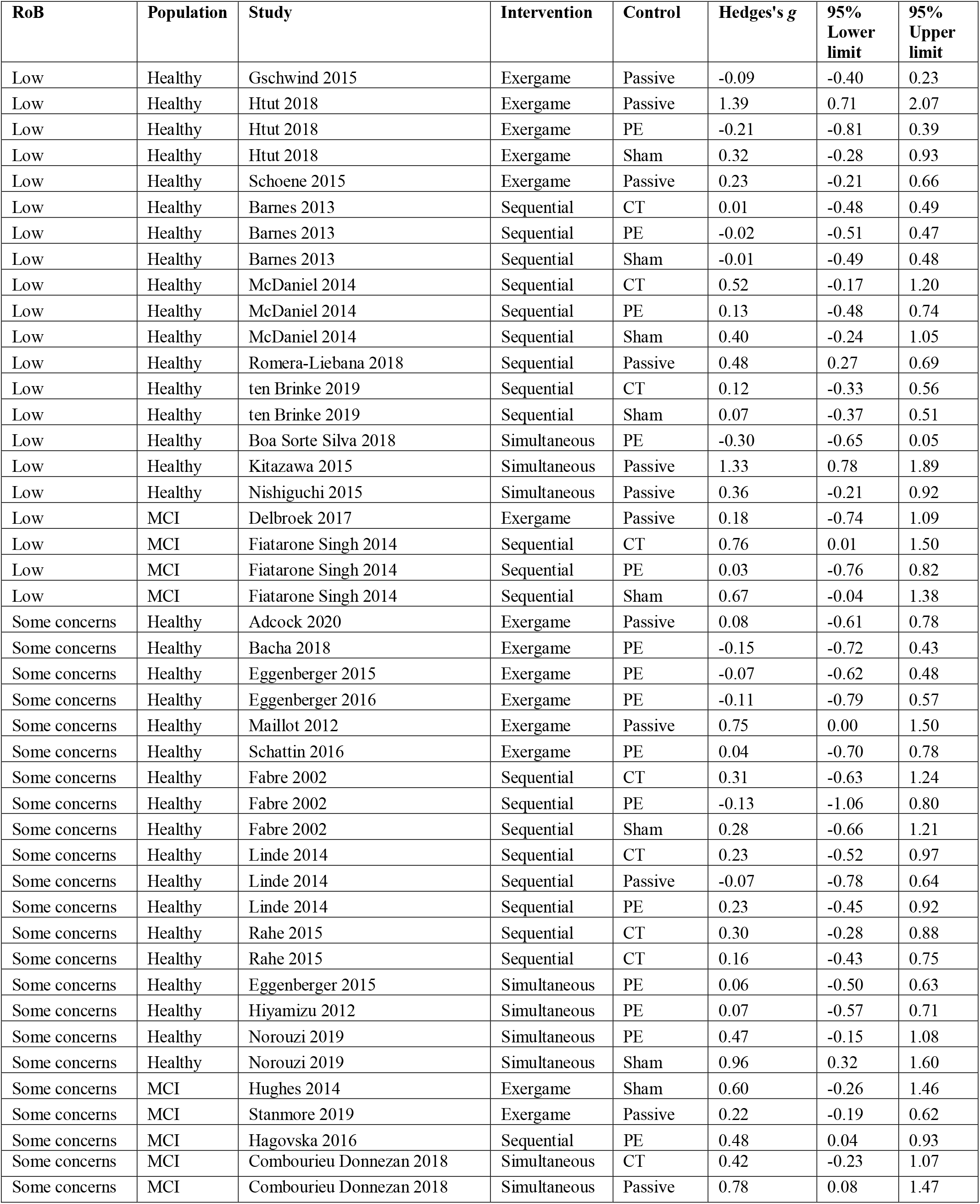
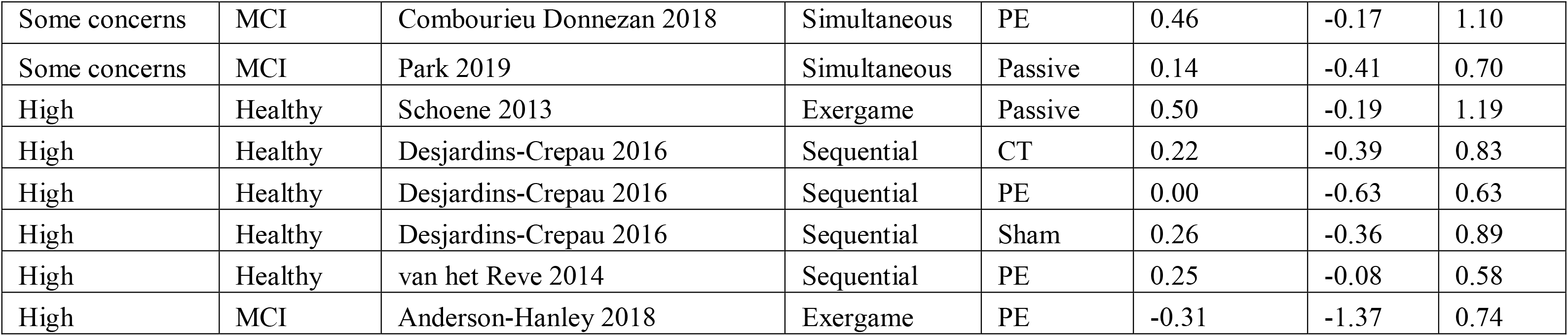

## Appendix H. Results of individual studies and comparisons: Overall psychosocial outcomes

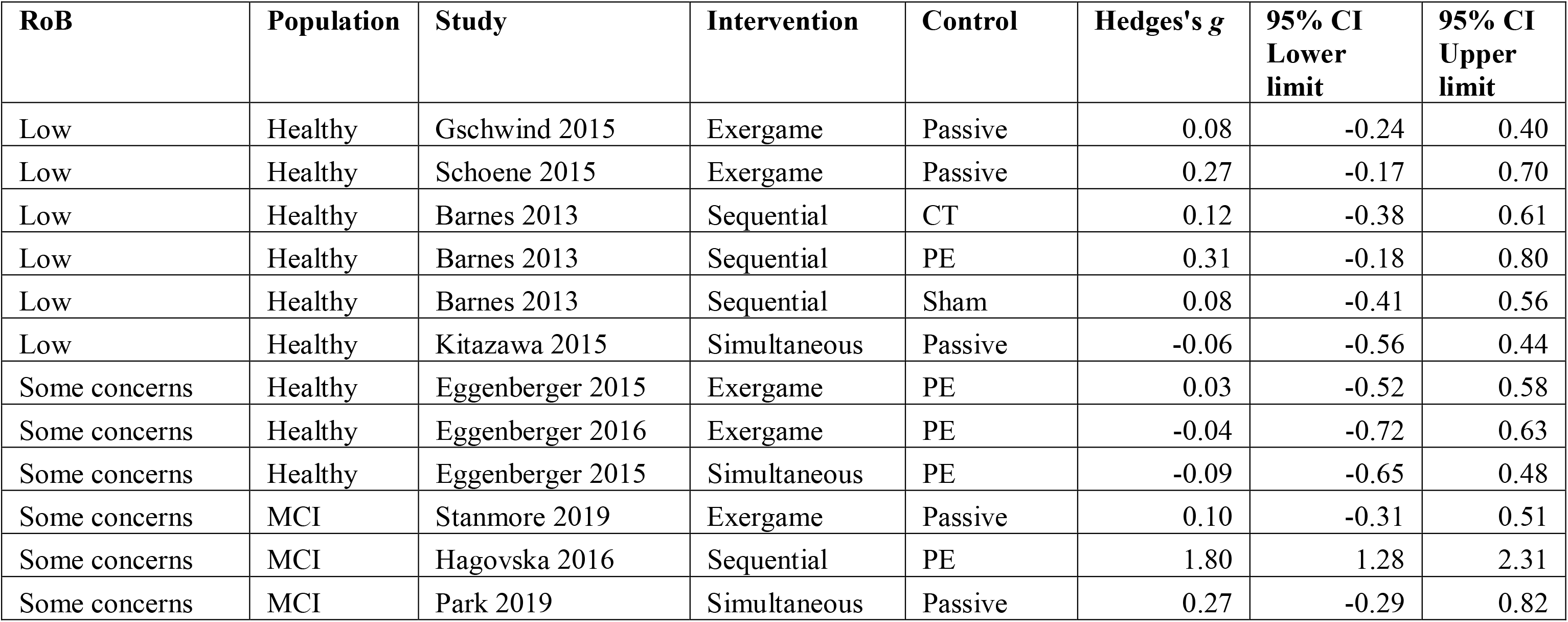

## Appendix I. Results of individual studies and comparisons: Dementia and Parkinson’s disease

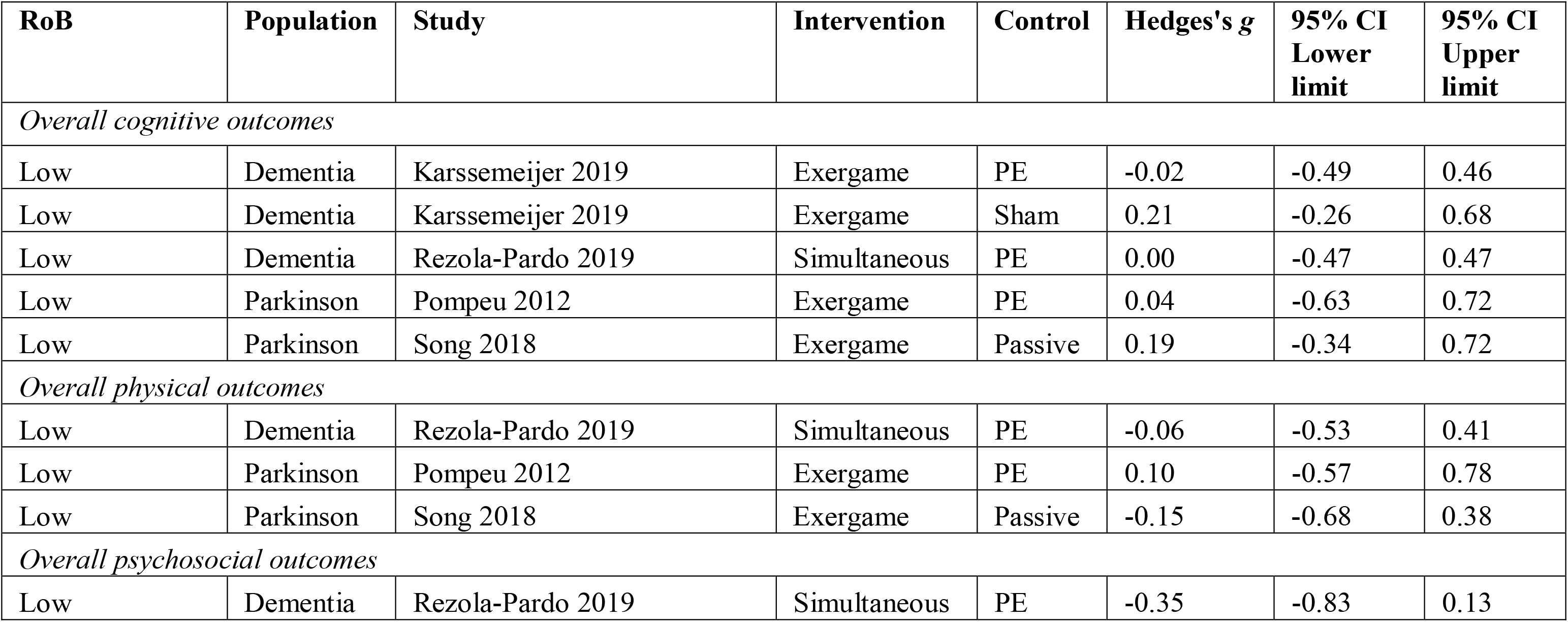

## Appendix J. Distribution of potential effect modifiers

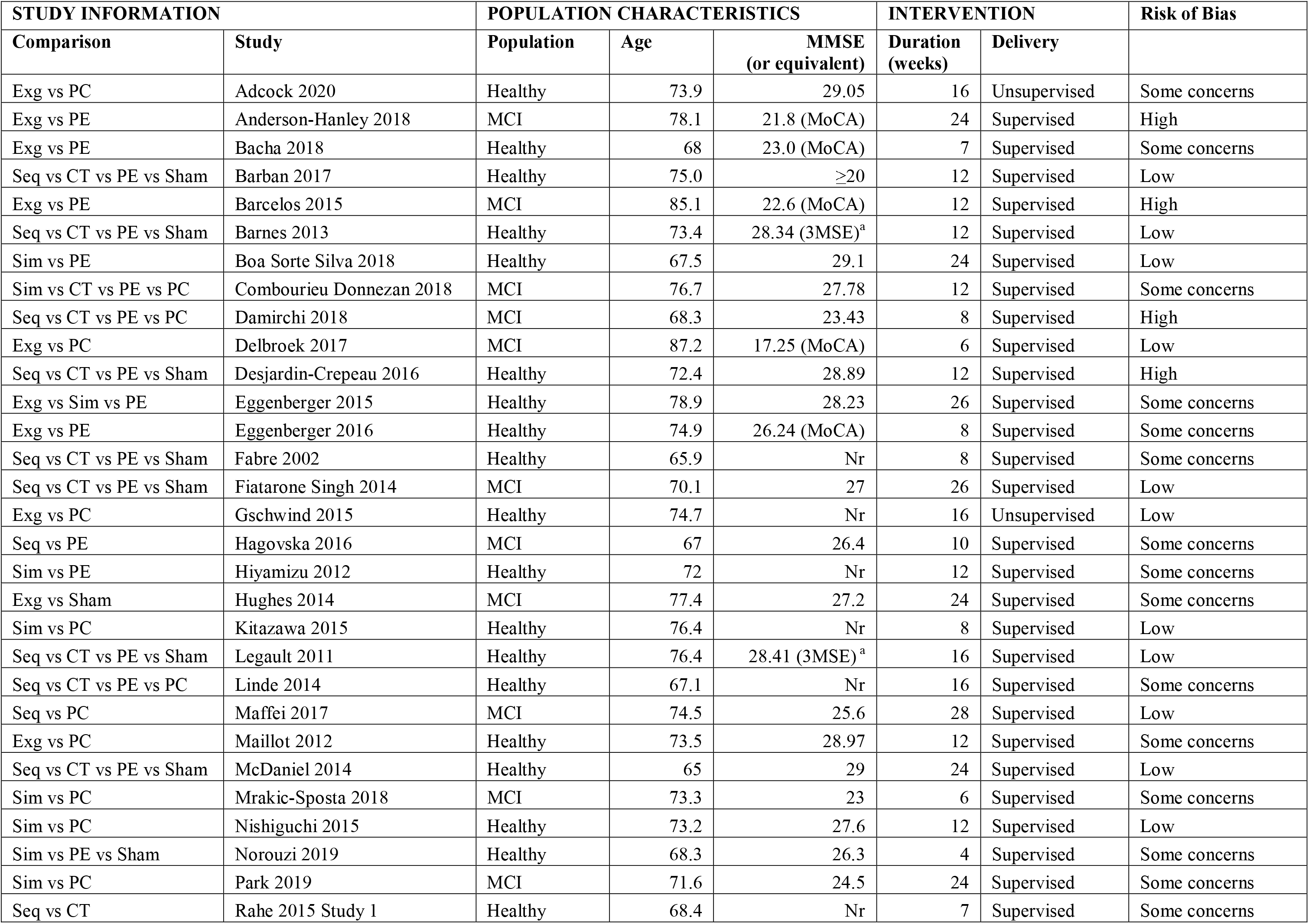
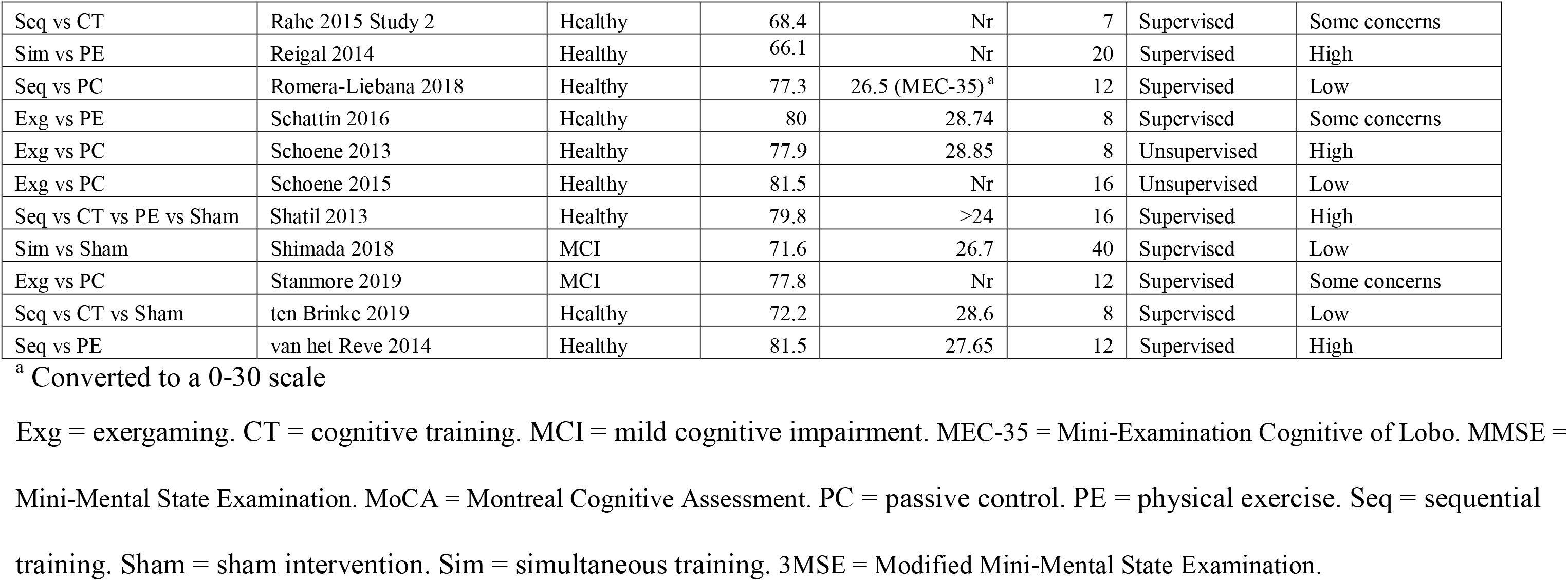

## Appendix K. CINeMA grading of the certainty of the evidence

We evaluated the certainty of the evidence for the network meta-analysis using the CINeMA web application (Papakonstantinou, Nikolakopoulou, Higgins, Egger, & Salanti, 2020), applying the following criteria:

### Within-study bias

The overall risk of bias assessment for each study was combined with the percentage contribution matrix to compute the percentage contribution for each comparison from studies with a low, some concerns and high risk of bias. The overall assessment for each comparison was based on the average risk of bias of the contributing studies.

### Reporting bias

No asymmetry was detected in the funnel plot and no significant association was found between study precision and effect size for cognitive or physical outcomes. Consequently, reporting bias was set as “undetected” for all comparisons.

### Indirectness

All of the included studies matched the study question and visual inspection of potential effect modifiers showed that these were similarly distributed across the network of included comparisons. However, four studies used unsupervised training, all of which compared exergaming with passive control. These studies were therefore downgraded to “moderate” indirectness. All other studies were set as “low” indirectness.

### Imprecision, heterogeneity and incoherence

We considered a clinically meaningful threshold for SMD to be 0.20.

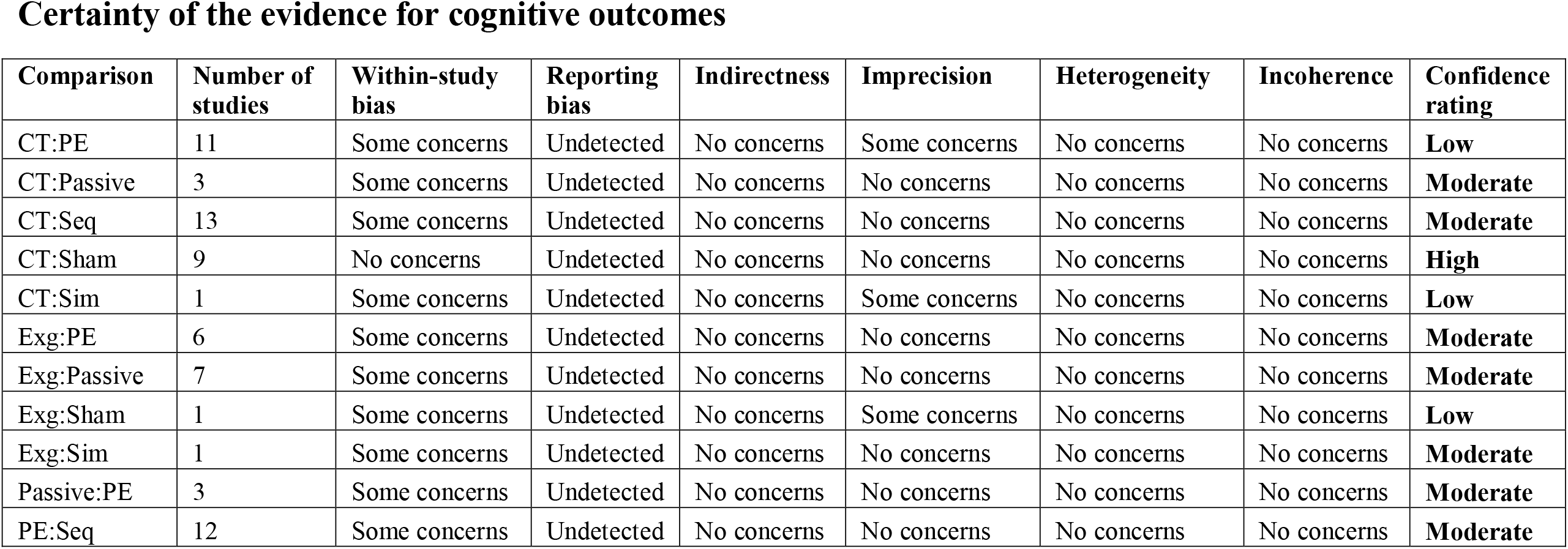
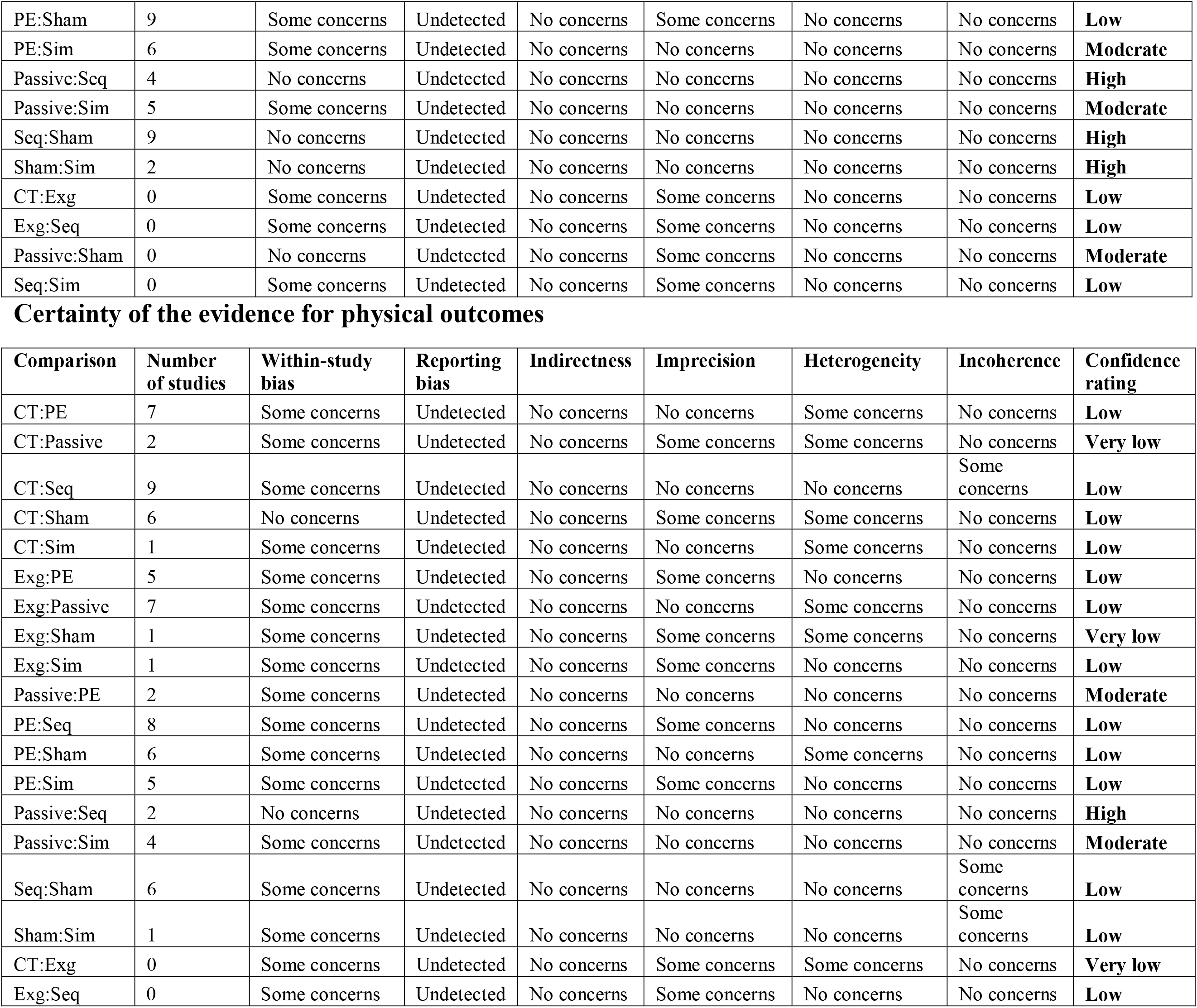
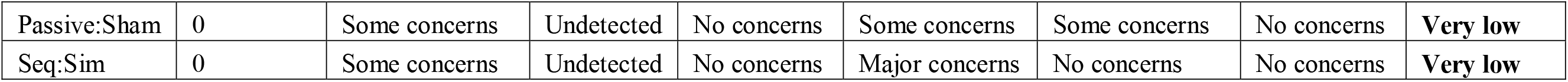

## Appendix L. Sensitivity analyses

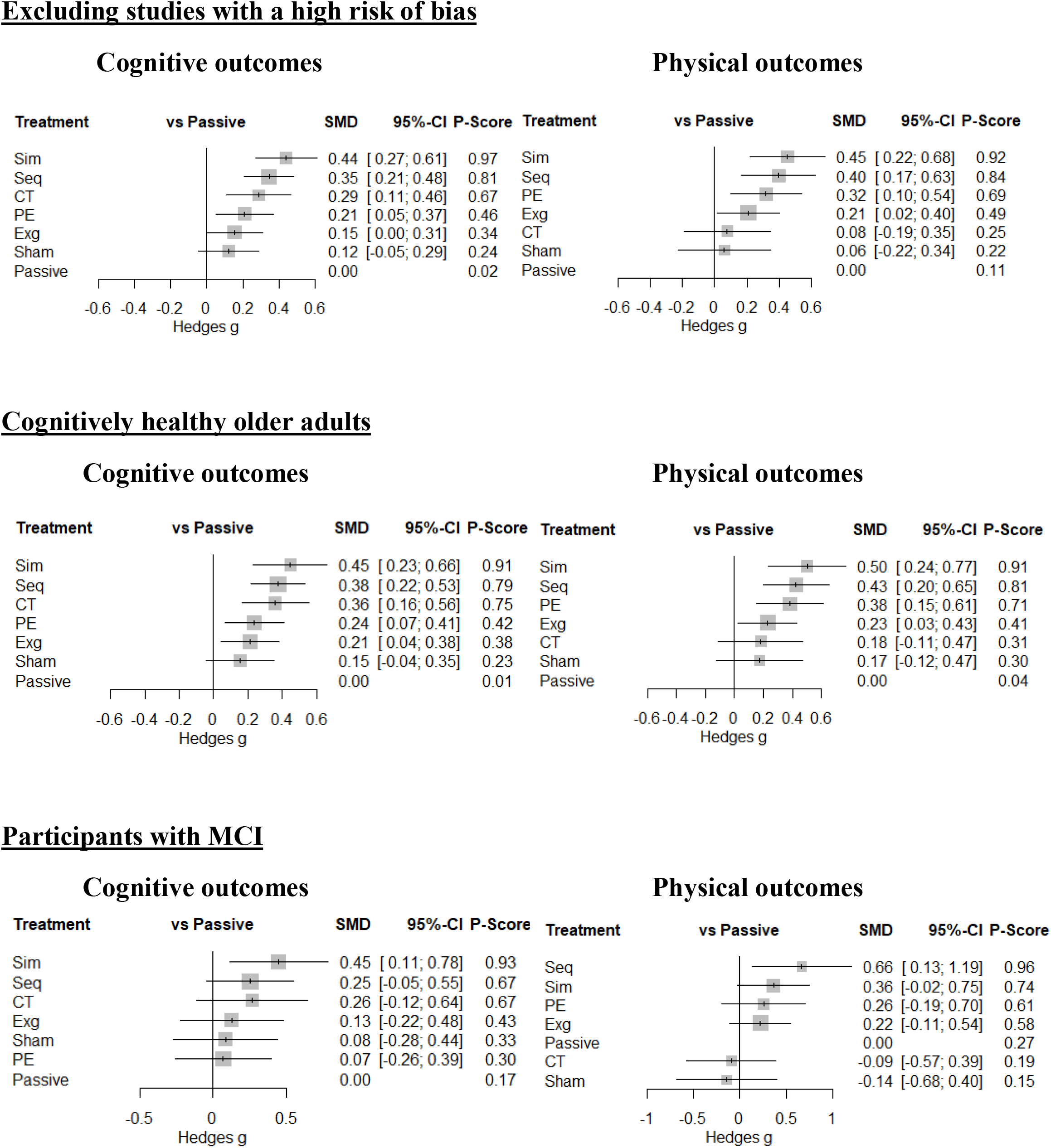

